# Interferon Beta-1α ring prophylaxis to reduce household transmission of SARS-CoV-2: the Containing Coronavirus Disease-19 randomized clinical trial

**DOI:** 10.1101/2022.06.13.22276369

**Authors:** José A. Castro-Rodriguez, Eleanor N. Fish, Tobi Kollmann, Carolina Iturriaga, Yuliya Karpievitch, Casey Shannon, Virginia Chen, Robert Balshaw, Samuel T. Montgomery, Joseph Ho, Rym Ben Othman, Radhouana Aniba, Francisca Gidi-Yunge, Lucy Hartnell, Guillermo Pérez-Mateluna, Marcela Urzúa, Scott Tebbutt, Diego García-Huidobro, Cecilia Perret, Arturo Borzutzky, Stephen M. Stick

**Author notes:** Corresponding author: Professor Stephen Stick.

## Abstract

**Importance:** Evidence suggests that early, robust type 1 interferon responses to SARS-CoV-2 are critical determinants for COVID-19 disease outcomes, accelerating viral clearance and limiting viral shedding.

**Objective:** We undertook a ring prophylaxis study to determine whether pegylated IFNβ-1α could reduce SARS-CoV-2 household transmission.

**Design:** A cluster randomized clinical trial of pegylated IFNβ-1α conducted in Santiago, Chile. Recruitment was conducted between December 4^th^ 2020, and 31^st^ May 2021, with the last follow-up completed June 29^th^ 2021.

**Setting:** The study was conducted across 341 households in the metropolitan area of Santiago, Chile.

**Participants:** Index cases were identified from databases of those with confirmed SARS-CoV-2 from COVID-19 clinics and emergency room visits in Santiago, Chile. 5,154 index cases were assessed for eligibility, 1,372 index cases were invited to participate, and 341 index cases and their household contacts (n = 831) were enrolled in the study.

**Intervention:** Households were cluster randomized to receive 125µg subcutaneous pegylated IFNβ-1α (n = 172 households, 607 participants), or standard care (n = 169 households, 565 participants).

**Main Outcome(s) and Measure(s):** Frequentist and Bayesian analyses were undertaken to determine the effects of treatment on (i) reducing viral shedding in index cases and (ii) reducing viral transmission to treatment-eligible household contacts. Four secondary outcomes were assessed including duration of viral shedding, effects on viral transmission and seroconversion, incidence of hospitalization, and incidence and severity of reported adverse events. A post-hoc ‘at risk population’ was defined as households where the index case was positive at the start of the study and there was at least one treatment eligible contact in a household who tested negative for SARS-CoV-2.

**Results:** In total, 1172 participants in 341 households underwent randomization, with 607 assigned to receive IFNβ-1α and 565 to standard care. Based on intention to treat and per protocol analyses, IFNβ-1α treatment was ineffective. However, in the ‘at risk’ population, the relative risk of infection was reduced by 23% in treated individuals and that there was a 95% probability that IFNβ-1α reduced household transmission

**Conclusions and Relevance:** Ring prophylaxis with IFNβ-1α reduces the probability of SARS-CoV-2 transmission within a household.

**Trial Registration:** Clinicaltrials.gov identifier: NCT04552379

## Introduction

The SARS-CoV-2 pandemic has claimed over six million lives. Despite the rapid development and deployment of vaccines in many countries, the number of new cases worldwide still exceeds 500,000 daily (https://covid19.who.int). With each wave of the pandemic, health systems have been challenged and the likelihood of emergence of mutant strains of the virus remains. Mutated strains may be more transmissible ^1,2^, cause more severe disease than the original pandemic strain of SARS-CoV-2 ^3^ and have the potential to evade available vaccines ^4,5^. Whilst widespread vaccination has had success limiting the trajectory of the pandemic, the emergence of the omicron variants demonstrated that even with mutations that appear to cause less severe disease ^6^, high transmission despite immunization could still result in significant pressures on health services ^7^.

The solution to halting any pandemic is ending community transmission. During the current pandemic, measures such as healthy hygiene, self-isolation when sick, physical-distancing and use of facemasks have all been effective ^8^. Moreover, expedited public health responses such as extensive contact-tracing, testing for infection and community lockdowns have all been effective methods to curb transmission ^9^. International and local border closures plus strict quarantine measures have reduced community transmission to zero for periods in countries such as Australia and New Zealand ^10,11^. However, in these countries and elsewhere, as restrictions are relaxed, localized outbreaks have occurred ^12^ that have required rapid, community-wide responses to again supress transmission. Importantly, these community constraints cause unprecedented civil disruption and come at enormous economic ^13,14^ and social costs ^15,16^.

Since the evolution of dominant SARS-CoV-2 virus cannot be easily predicted ^17^, there remains a need to identify interventions that can be rapidly deployed should highly pathogenic strains emerge despite high levels of community immunization. Furthermore, preparations for the next pandemic must include strategies to limit the potential for infection and transmission on first contact with pathogenic respiratory viruses.

One of the many therapeutic approaches investigated that appeared clinically useful early in the course of the pandemic was treatment with type 1 interferons (IFNs). Randomized, controlled studies suggest that IFNs offer clinical benefits in moderate ^18^ and severe disease ^19,20^ }, reduce the duration of viral shedding ^21^ and prevent infection in front-line hospital workers ^22^. Nonetheless, IFNs are not generally recommended for treatment of proven cases of Covid-19 ^23^.

Since IFNs are sentinel innate immune signalling molecules produced early after first contact with viral pathogens ^24–26^, we postulated that prophylactic IFN treatment might reduce susceptibility to infection of uninfected contacts of individuals with SARS-CoV-2 infection. Such post-exposure prophylaxis could provide non-specific, antiviral protection to curb episodic viral outbreaks ^27^, help suppress community transmission, even in vaccinated populations, and therefore reduce the risk of emergence of dangerous mutations ^4,28^.

We therefore undertook a cluster, randomized, controlled study of sub-cutaneous pegylated IFNβ-1α (Plegridy. Biogen Inc, Cambridge MA) administration to determine whether IFNβ-1α therapy given to index cases and household contacts can reduce transmission of SARS-CoV-2.

## Methods

The Containing Coronavirus Disease-19 (ConCorD-19) trial was a prospective, cluster, randomized trial of subcutaneous (sc) administration of pegylated IFNβ-1α (“IFNβ-1α”) versus standard care (control) ^29^. Each household of an index case was randomized to either the IFNβ-1α treatment or the standard care control arm. The study was approved by the Institutional Review Board of the Pontificia Universidad Católica de Chile and was registered with clinicaltrials.gov (NCT04552379). All participants provided written informed consent.

### Trial population

Index cases were identified from databases of those with confirmed SARS-CoV-2 from COVID-19 clinics and emergency room visits in Santiago, Chile. Households were contacted by telephone to determine eligibility prior to enrolment. Individuals aged between 18 and 80 years who met inclusion and exclusion criteria were deemed as ‘eligible’ contacts with households only included if there was at least one eligible contact ^29^. Enrolment and the first doses of IFNβ-1α - if in the intervention arm-had to be within 72 hours of the positive identification of SARS-CoV-2 by polymerase chain reaction (PCR) in index cases. A household was excluded if the index case had been in complete self-quarantine from other household members during the 48 hours prior to the diagnosis of SARS-CoV-2 infection ^29^. Household characteristics were captured consistent with recommendations of the World Health Organization for assessing household transmission ^30^. All participants implemented quarantine measures as mandated by local authorities and maintained a daily symptom diary which was collected and reviewed at each study visit by the study team (see online supplementary material). At any time during the trial, if a contact participant developed symptoms suggestive of COVID-19, a nasopharyngeal swab was taken by a member of a mobile health team for virus PCR testing in an accredited SARS-CoV-2 diagnostic laboratory. Index cases were instructed to remain in isolation / quarantine for 11 days from onset of symptoms or, if asymptomatic, 11 days from the sample collection date that resulted in the COVID-19 diagnosis. Household contacts remained in isolation / quarantine for 11 days from date of the sample resulting in the diagnosis of the index case or of a newly diagnosed household member as per recommendations/rulings by local authorities.

### Intervention

A mobile health team conducted home visits of all participant households on study days 1 (enrolment), 6, 11, 16 and 29. Index cases and eligible household contacts in the IFNβ-1α arm received three subcutaneous doses of IFNβ-1α (125µg/0.5ml x 0.5ml) on study days 1, 6 and 11. Ineligible contacts in the IFNβ-1α arm, as well as index cases and all household contacts in the SOC arm, received standard care. All participating households received information regarding hygiene, isolation, social distancing and wearing of facemasks as per public health advice at the time of enrolment. The IFNβ-1α injection was given by a trained member of a mobile health team and participants were recommended to take paracetamol (1000mg, 6 hourly) commencing at the same time as the IFNβ-1α for up to twenty-four hours, in order to mitigate predictable flu-like symptoms ^31^.

### Outcomes

The primary outcomes were (i) the proportion of index cases shedding SARS-CoV-2 at study day 11 in the IFNβ-1α compared to control arm and (ii) the proportion of household contacts shedding SARS-CoV-2 at day 11, in the IFNβ-1α compared to the control arm. Secondary and exploratory outcomes are listed in Supplementary Methods (Supplementary Table 1). Shedding was determined by the presence of SARS-CoV-2 by PCR in saliva collected on days 1, 6, 11, 16, 21, and 29 (see Supplementary Methods). A SARS-CoV-2 PCR Ct value ≥40 was considered negative. Viral load was estimated from the Ct value using a standard algorithm (Supplementary Figure 1

### Biospecimen collections

The full schedule of biospecimen collection is provided in the supplementary methods (Supplementary Tables 2 and 3. All consenting, non-eligible household contacts also provided biospecimens according to the schedule collection for non-eligible household contacts.

### Adverse events

These were classified in accordance with Good Clinical Practice ^32^, as non-serious or serious and as related or unrelated to the trial medication.

### Statistical considerations

The study was designed at the start of the pandemic and there were few data to guide sample size calculations. We therefore planned both frequentist inference and Bayesian analyses based on assumptions from the available data.

Households were randomized as individual clusters using a minimization technique (biased coin, P=0.7) in order to achieve balance between treatment arms, stratified by the number of people within the household ^33^.

Sample size calculations assumed, α=0.025 and power 90%, a zero correlation between outcome measures and that the household-wise, type 1 error rate would be below 0.05 and power above 80%. The estimated average household size was 4, based on census data ^34^.

Primary outcomes:

i. SARS-CoV-2 shedding at study day 11 by index cases: Data from Wuhan suggested that the proportion of untreated index cases still shedding virus on study day 11 would be ~85% ^35^. Based on a two tailed Fisher’s exact test, a sample size of 278 index cases (139 per arm) would have 90% power to detect a difference in the proportion of individuals shedding virus at study day 11, if the proportion in the IFNβ-1α arm was 65%.
ii. SARS-CoV-2 shedding at study day 11 by household contacts: We estimated the secondary infection rate (transmission within the household) where there is an untreated index case would be 28% ^36,37^. Based on a stratified Cochran-Mantel-Haenszel test (two-tailed with intra-class correlation of 0.15), we estimated that a sample of 278 households (834 eligible household contacts) would have >90% power, at alpha 0.025, to detect an odds ratio of 0.5 for a reduction in transmission to a treated household contact.

Based on these assumptions and estimates, we aimed to recruit 310 households (allowing for a 10% drop-out rate) and 930 eligible household contacts.

We undertook 3 separate sets of analyses:

A. Standard frequentist analyses based on the primary outcomes as stated in the protocol and described in the Statistical Analysis Plan (available online) and included intention to treat and per protocol approaches.
B. Exploratory frequentist analyses based on a modified dataset that accounted for biological implausibility and effects that might only be associated with active treatment (Figure 1 and Supplementary Table 1 for further explanations). Both frequentist analyses fitted linear mixed effects models using Ime4 ^38^ (see Statistical Analysis Plan).
C. Exploratory Bayesian analyses using the same dataset as the exploratory frequentist analysis. Detailed methods are provided in the supplementary methods. Briefly, a generalized linear model with mixed effects was developed to estimate the probability of infection that is influenced by explanatory variables for each contact case. A stan_glmer with binomial logit function in rstanarm R package was used ^39,40^.

**Figure 1:**
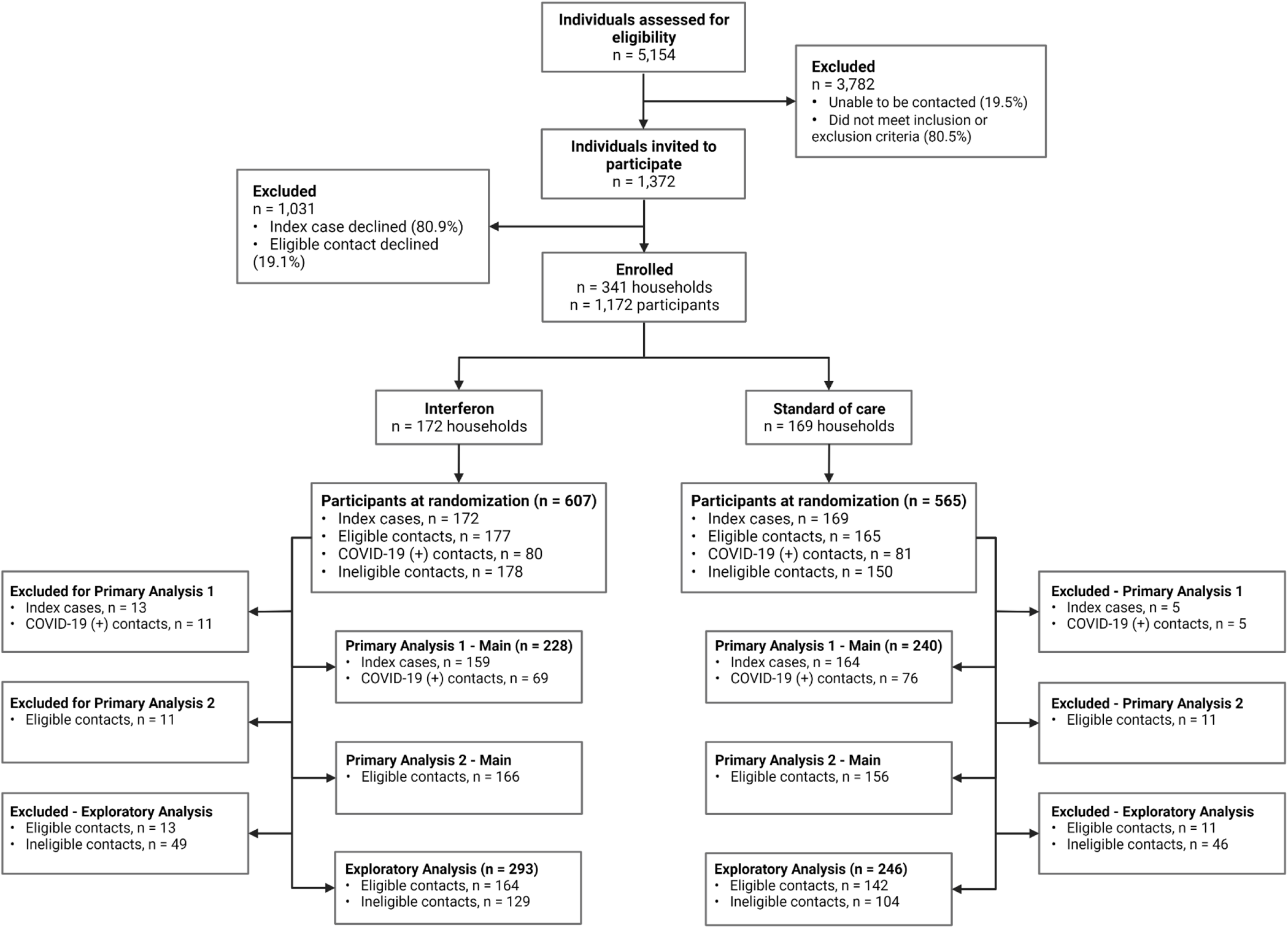
CONSORT diagram describing participant screening, enrolment, randomisation, and analysis.

The frequentist approach allowed us to estimate the effects of IFNβ-1α treatment on the risk of an *individual* becoming infected. Whereas the Bayesian analysis allowed us to determine whether the ring prophylaxis strategy using IFNβ-1α reduces the probability of transmission within the household of an infected index case.

With regard to the filtered datasets, we observed, after locking the database, that a number of index cases had negative PCR results on the day of recruitment despite a previous positive diagnostic PCR test. In the exploratory analyses we therefore excluded households where the index case was unlikely to transmit virus because they were not shedding virus on days 1 and day 6 of the study. We also excluded households where an eligible contact was already positive at recruitment and there were no other eligible negative cases in the household. We refer to the modified dataset as the population “at risk”. We used the population at risk data to better understand the effects of IFNβ-1α treatment on transmission of virus within households.

When undertaking these exploratory analyses, we considered effects during two time periods. Days 1-11 (Period 1) i.e., the treatment and isolation period when eligible contacts in the IFNβ-1α arm were likely to have elevated IFN levels based on the known pharmacokinetics of pegylated IFNβ-1α and days 12-29 (Period 2) the post-treatment and isolation period when biological effects of pegylated IFNβ-1α were likely to be waning in treated individuals. Although samples were not obtained on days 12-15, we assumed that anyone who became infected during this period would likely still to have a positive PCR on day 16.

## Results

Recruitment, participation, and completion data are shown in Figure 1. Participant demographic and basic household characteristics data are shown in Table 1.

**Table 1:**
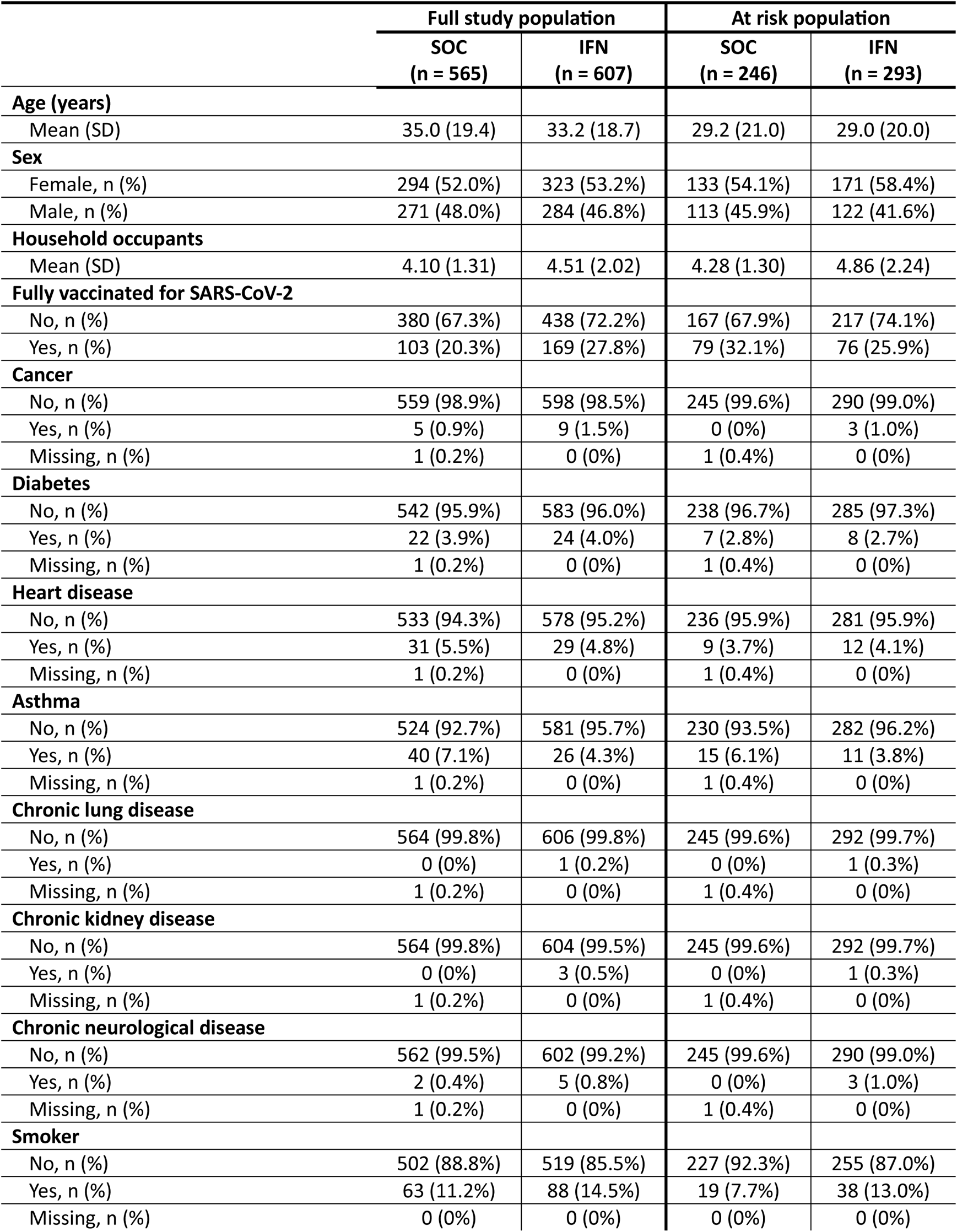
Full study population demographics at baseline, and at risk population demographics at baseline.

Between December 2020 and May 2021, three-hundred and forty-one households were enrolled and randomized, of which 137 (IFNβ-1α arm) and 151 (control arm) completed the study. Of the 1172 individuals randomized (IFNβ-1α arm = 607; Controls = 565), 53 individuals withdrew from the study, of which 15 were index cases; 35 (14 index cases) in the IFNβ-1α arm and 18 (1 index case) in the control arm. The reasons for withdrawal are summarized in the supplementary results (Supplementary Table 4). One index case withdrew before being allocated to a treatment arm.

Eighty-two households where the index case had a negative salivary PCR on Day 1 and 6, or where there were no eligible contacts who tested negative at recruitment, were excluded from the exploratory analyses. The remaining households were considered as the ‘at risk’ population. (Figure 1).

a. *Primary analyses* The outcomes for the intention to treat (ITT) and per protocol (PP) frequentist analyses were similar (Supplementary Results). There was no effect of IFNβ-1α treatment on duration of viral shedding in those treated (index cases or infected contacts) vs. controls (IC-INF: treatment OR = 0.979, 95%CI = 0.647 to 1.479); IC-ITT: treatment OR = 1.106, 95%CI = 0.657 to 1.863); IC-PP: treatment OR = 1.062, 95%CI = 0.622 to 1.810); Figure 2A). The observed reduction in transmission to household contacts associated with IFNbeta-1alpha by day 11 was not significant (EHC-ITT: treatment OR = 0.582, 95%CI = 0.271 to 1.247; HC: treatment OR = 0.577, 95%CI = 0.287 to 1.160; EHC-PP: treatment OR = 0.589, 95%CI = 0.272to 1.276; Figure 2B). Vaccination status did not affect these outcomes (Supplementary results).
b. *Exploratory frequentist analysis of ‘At Risk Population*.’ Of the at risk population, 33/164 (20%) in the IFNβ-1α arm became infected by day 29 of the study compared to 37/142 (26%) in the control arm (relative risk reduction in the IFN arm = 23%). The majority of the risk reduction occurred during the active treatment phase of the study (days 1 to 11). Treatment with IFNβ-1α was associated with a significant reduction in the odds of a positive saliva PCR for all household contacts compared to standard of care on Day 11 (p = 0.033; OR = 0.550, 95% CI = 0.364 to 0.989). The treatment effect was not significant in the second period (days 12-29) was not significant (OR = 0.968, 95% CI = 0.405 to 2.840) (Figure 2C)
c. *Exploratory Bayesian analysis:* Overall, there was a 95% probability of infection reduction within a household by IFNβ-1α treatment and the credible interval for the reduction in transmission probability was in the order of 0.9 to 15.9%. During the active treatment period (days 1-11), there was a significant reduction in the odds of transmission (Bayesian analysis (OR = 0.32, 95% Credible Interval = 0.11 to 0.83). In contrast, in Period 2 (day 12-29), the 95% credible interval of *Beta* coefficients for the treatment group includes zero and therefore treatment was not effective in this period (Figure 2C, Supplementary Figure 2). The effect of IFNβ-1α on transmission was independent of household size (Supplementary Figure 3)

**Figure 2:**
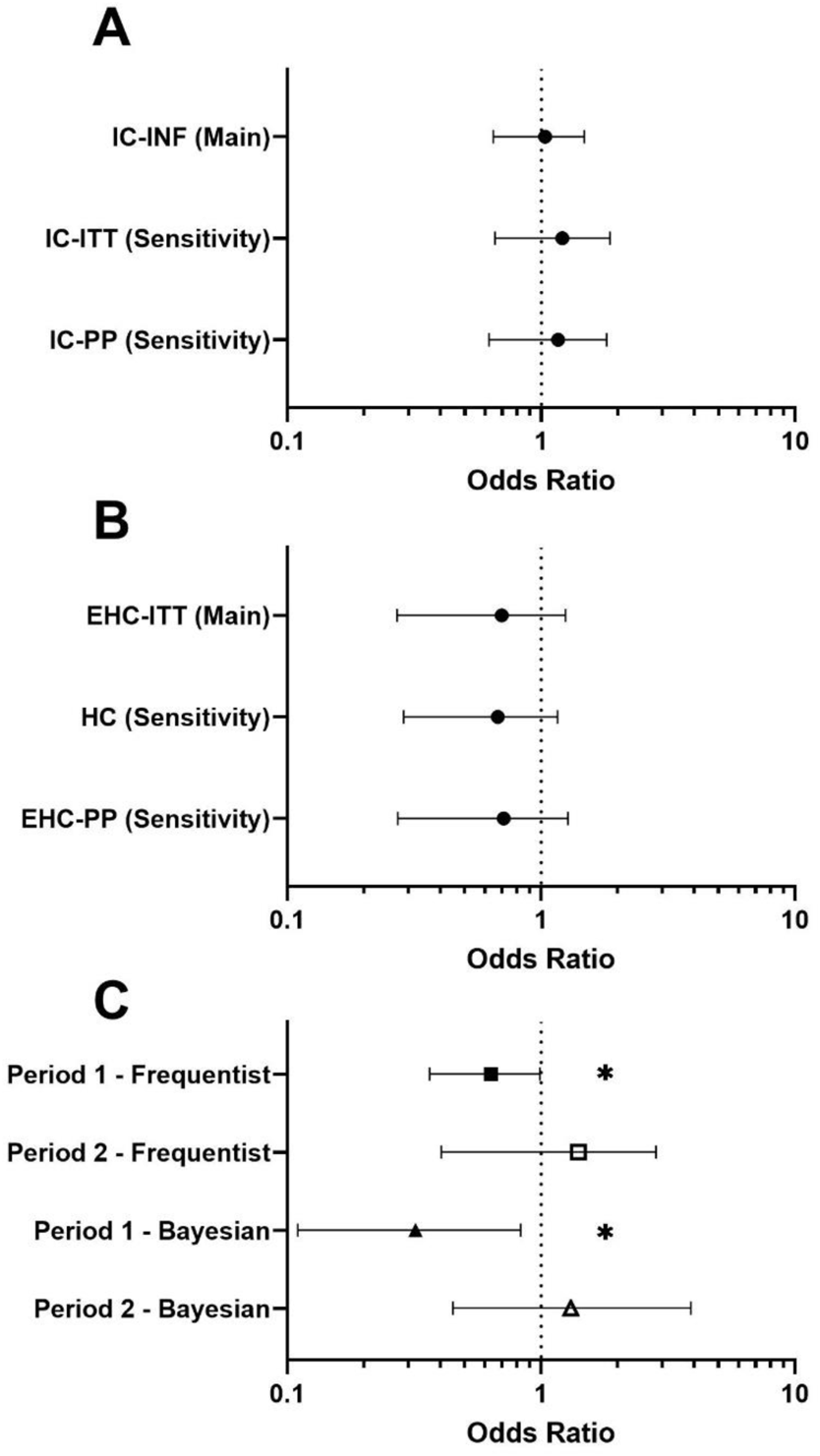
Forrest plots describing primary and exploratory study outcomes. **(A)** odds ratios for index case populations in Primary Analysis 1 and associated sensitivity analyses, to compare the probability of being SARS-COV-2 negative vs positive on study day 11 in either treatment or standard of care arms. **(B)** odds ratios for household contact populations in Primary Analysis 2 and associated sensitivity analyses, to compare the probability of household contacts being SARS-CoV-2 negative vs positive on study day 11 in either treatment of standard of care arms. **(C)** odds ratios for the exploratory frequentist and Bayesian analysis, to determine if IFN is associated with a reduction in the number of positive COVID-19 tests in household contacts across study period 1 (days 1-11) and study period 2 (days 12-29). Treatment with IFN is associated with a significant reduction in the odds of a positive COVID-19 test for all household contacts compared to standard of care; frequentist analysis (p = 0.033; OR = 0.550, 95% CI = 0.364 to 0.989), Bayesian analysis (OR = 0.32, 95% CrI = 0.11 to 0.83). IC-INF = Infected case population (main analysis), IC-ITT = Index case – intention to treat population (sensitivity analysis 7.1.3.2), IC-PP = Index case – per protocol population (sensitivity analysis 7.1.3.3). EHC-ITT = Treatment-eligible household contact – intention to treat population (main analysis), HC = Household contact (eligible + non-eligible contacts) population (sensitivity analysis 7.2.3.2), EHC-PP = Treatment-eligible household contact = per protocol population (sensitivity analysis 7.2.3.3). * p < 0.05.

To determine the effect of IFNβ-1α treatment on viral shedding in those who became infected, we examined the viral load trajectories in household contacts who became PCR positive after day 1. We did not observe any difference in viral load trajectory in those treated with IFNβ-1α compared to controls (Figure 3A). Furthermore, there was no difference in peak viral load between treated household contacts and controls who became infected (Figure 3B).

**Figure 3:**
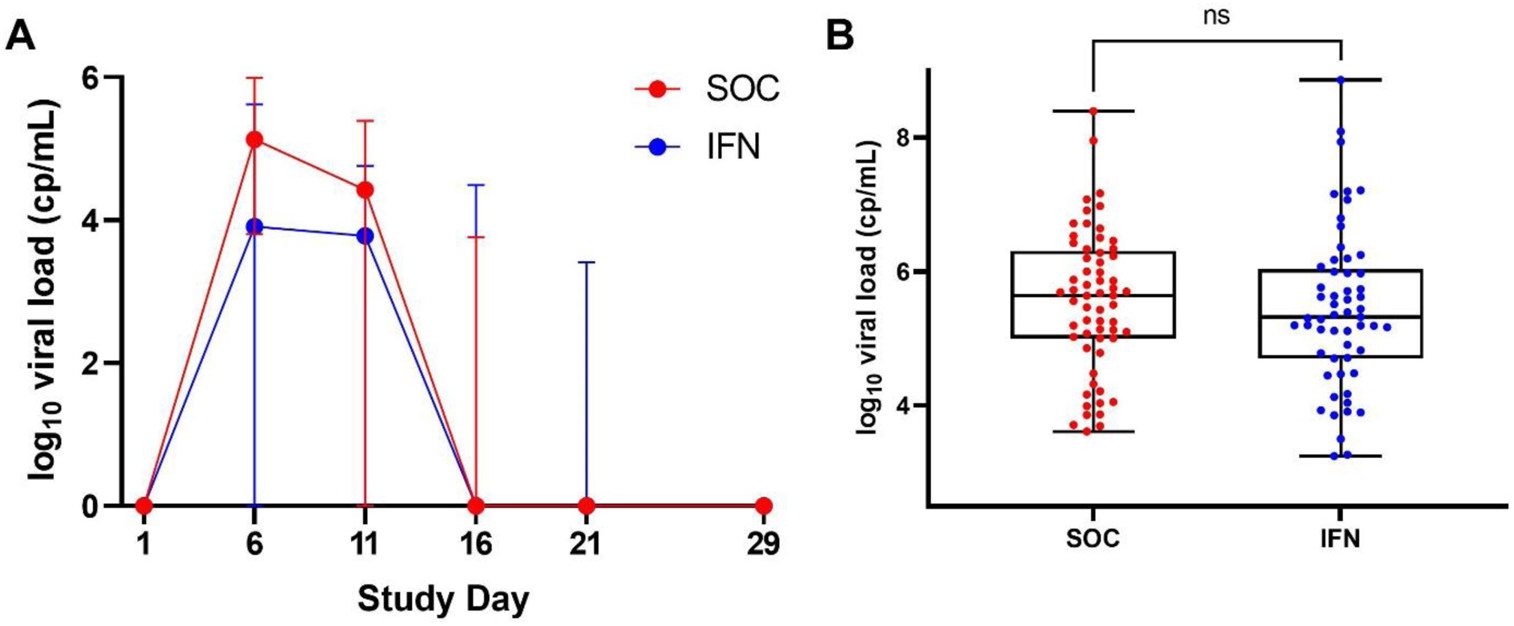
Log_10_ viral load in study participants who test positive for COVID-19 after visit 1. **(A)** No significant difference in median viral load at each study visit between SOC and IFN arms. **(B)** No significant difference in peak viral load between SOC and IFN arms. SOC = standard of care arm, IFN = interferon arm, cp/mL = copies/mL. n = 57 (IFN) & n = 61 (SOC) participants per group.

There were fifty-eight serious adverse events in index cases, twenty-seven hospitalizations due to COVID-19 (25 in the treatment and 32 in the control arm) and was one death in the standard care arm due to COVID-19 in an individual with significant co-morbidities. Twenty-six household contacts of index cases were hospitalized, 10 in the treatment arm and 16 in the control arm, the difference was not statistically significant.

## Discussion

This prospective, household cluster randomized, ring prophylaxis trial demonstrated that IFNβ-1α may reduce household transmission of SARS-CoV-19. Pegylated IFNβ-1α is an FDA approved therapy for multiple sclerosis for which the pharmacokinetics and safety profile are well-characterized ^41^. The use of this formulation allowed us to predict the likely duration of activity of the three-dose regimen in order to cover the period of peak transmissibility of the virus. Using a mobile medical team to administer doses at home optimized adherence to therapy and allowed reasons for non-adherence and withdrawal to be accurately documented.

Although the intention to treat and per protocol analyses failed to demonstrate a significant effect on primary outcomes, these analyses failed to account for index cases not shedding virus at randomization nor an absence of eligible contacts within a household. Using biologically plausible filters to define an at risk population provided a better understanding of the effects of IFNβ-1α than intention to treat or per protocol analyses. These exploratory analyses that took account of the likelihood that transmission could occur within a household provided a pragmatic assessment of the effects of IFNβ-1α therapy when given to household contacts as prophylaxis.

The frequentist analysis of the at risk populations indicated that *individuals* treated with IFN were less likely to become infected than untreated individuals when exposed to virus with a relative risk reduction of 23%. The Bayesian analysis revealed that the probability was 95% that transmission was reduced in *households* randomized to IFNβ-1α therapy. This effect of IFNβ-1α on household transmission was only significant during the active treatment period. This highlights the physiological importance of IFNβ-1α as a sentinel molecule ^24^ and provides further evidence for post-exposure prophylaxis with IFNβ-1α as an antiviral strategy.

To our knowledge, only one previous study assessed ring prophylaxis in COVID-19. Labhardt et al observed that a combination of lopinavir/ritonavir for 5 days as post-exposure prophylaxis was not effective at preventing infection in close contacts of index cases ^42^.

The overall viral load trajectory we observed was very similar to that observed in a laboratory human challenge experiment using wild-type SARS-CoV-2 virus in healthy volunteers ^43^. Treatment with IFNβ-1α had no effect on viral load trajectory; therefore, reduced viral shedding by infected individuals is unlikely to explain the protective effect on household transmission of IFNβ-1α treatment that we report. Given that treatment affected the probability of transmission only in the active treatment phase and appears unrelated to viral load, we speculate that the observed effects of IFNβ-1α were direct through protection of the at risk, exposed individual rather than indirectly though effects on the index case. The ConCoRD-19 biorepository will allow further examination of the mechanisms of action of IFNβ-1α on resistance to infection.

The study was undertaken prior to the emergence of the omicron strain of virus (Supplementary Figure 4). We calculated the sample size based upon known household transmission characteristics of the alpha strain which was the dominant strain of the virus early in the pandemic. Simple hygiene measures and quarantine of affected individuals within households could have contributed to lower rates of transmission than expected. Together with a smaller number of eligible household contacts than anticipated from census data, these factors may have reduced the power of the study for the primary outcomes. The effect size observed in this study could be greater for variants with higher transmissibility such as the Omicron strains that are now the dominant worldwide.

Nonetheless, the beneficial biological effect of IFNβ-1α treatment was evident in the at risk population and therefore, post-exposure or ring prophylaxis with IFNβ-1α warrants further investigation, particularly if additional virulent strains of SARS-CoV-2 emerge and for future pandemic virus preparedness.

In summary, although intention to treat and per protocol analyses failed to demonstrate significant effects on primary outcomes, in a biologically defined, at risk population, the relative risk of infection for an individual treated with IFNβ-1α was reduced by 23.0%. Moreover, this is the first study to demonstrate ring prophylaxis with IFNβ-1α significantly reduces the probability of household transmission of SARS-CoV-2. The effect of IFNβ-1α was most obvious during the active phase of treatment. In situations where there is emergence of a highly pathogenic and transmissible mutant strain of SARS-CoV-2 despite high vaccination rates, or when there is a new viral pandemic, IFNβ-1α ring prophylaxis could be considered to reduce transmission in households or amongst critical workers and vulnerable communities. Ring prophylaxis with therapies that can interrupt transmission appears to be an effective strategy for highly contagious respiratory viruses.

## Authorship

Authors Castro-Rodriguez; Fish; Kollmann; Iturriaga, Karpievitch and Shannon contributed equally to the conduct of the study and manuscript preparation as first authors. Authors Tebbutt, Diego García-Huidobro, Perret, Borzutsky and Stick contributed equally to the conduct of the study and manuscript preparation as senior authors.

## Data Availability

All data produced in the present study are available upon reasonable request to the authors

## Acknowledgements

Matthew Cooper (Telethon Kids Institute). Advice regarding protocol and establishment of randomization schedule. Alexia Foti (Telethon Kids Institute). Collating, reviewing, and editing documents. Zsuzsanna Hollander (PROOF Centre, Vancouver). Statistical advice and statistical analysis plan review. Nat Eiffler (Telethon Kids Institute). Project management, protocol development, data management. Jessica Meyer (University of Rochester). Assisting collation and formatting of tables and figures.

## Supplementary Materials

## Supplementary Methods

### Real-time Quantitative Polymerase Chain Reaction (RT-qPCR) for SARS-CoV-2

RNA extraction and RT-qPCR for SARS-CoV-2 detection was performed at the Infectiology and Molecular Virology Laboratory, Red Salud UC Christus, Santiago, Chile. RNA was extracted in parallel from saliva using the MagBind RNA extraction kit (#GN7101907, Maccura Biotechnology, Rockville, MD, USA) with the Auto-Pure 32A Nucleic Purification System (Maccura Biotechnology, Rockville, MD, USA), and using the RNA/DNA Purification Magnetic Bead Kit (#DA0630, DaAnGene, Guangzhou, CN) with the Smart32 Nucleic Acid Extraction Instrument (DaAnGene, Guangzhou, CN) according to manufacturer specifications. Real-time PCR amplification was performed using the LightMix Modular Wuhan CoV RdRP-gene kit (#53-0777-96, TIB MolBiol, Roche, Berlin, DE) targeting the RNA-dependent RNA polymerase (RdRP) gene, targeting SARS-COV-2, but not other SARS-like viruses (CoVNL63, CoV229E, HKE, OC43, MERS), with a positive control containing three separate diagnostic targets (E gene, N gene, RdRP). The reverse transcription and PCR processes occur in a single step in a real-time thermocycler (Lightcycler 480 II, Roche, Berlin, DE) with an analytical sensitivity of 10.6 copies per reaction. A cycle threshold (Ct) value of ≤ 37 was considered positive for SARS-CoV-2.

### Viral load calculation

The viral load of SARS-CoV-2 detected in saliva was calculated using a standard curve. A commercial SARS-CoV-2 standard (#COV019, Exact Diagnostics, Fort Worth, TX, USA) at a concentration of 200,000 copies per mL (cp/mL) was extracted in triplicate using the RNA/DNA Purification Magnetic Bead Kit (#DA0630, DaAnGene, Guangzhou, CN) with the Smart32 Nucleic Acid Extraction Instrument (DaAnGene, Guangzhou, CN) according to manufacturer specifications. Following extraction, serial dilutions (1:10; 200,000cp/mL to 20cp/mL & 1:5; 200,000cp/mL to 64cp/mL) were used in triplicate for reverse transcription and RT-qPCR using the LightMix Modular Wuhan CoV RdRP-gene kit (#53-0777-96, TIB MolBiol, Roche, Berlin, DE) with the Lightcycler 480 II thermocycler (Roche, Berlin, DE), resulting in 9 replicates at each concentration. SARS-CoV-2 was detectable in all nine replicates at 200,000cp/mL, 40,000cp/mL, 20,000cp/mL, 8,000cp/mL, 2000cp/mL, and 1,600cp/mL, and a standard curve was generated using the mean Ct values for these concentrations (Supplementary Figure 1) to calculate SARS-CoV-2 viral load in samples of unknown titre, resulting in a lower limit of quantification of 1,600cp/mL.

### Populations used for analyses

Participants were excluded for analysis as outlined in the statistical analysis plan, or as outlined in the exploratory analysis (Supplementary Appendix B). Total households and participants excluded and used in each population are described in Supplementary Table 6. Briefly, participants were excluded from the populations used in the primary analyses (IC-INF, IC-ITT, EHC-ITT, HC) if the study visit was not performed, or was performed outside the +/– 1 day window either side of the scheduled visit date (Statistical Analysis Plan 6.5). Participants who didn’t complete the full course of the treatment were excluded from the Per Protocol populations used in the primary analyses (IC-PP, EHC-PP). In the exploratory analysis, whole households were excluded if the index case was SARS-CoV-2 negative via salivary PCR at study days 1 & 6, and if there were no SARS-CoV-2 negative eligible or ineligible contacts in the household at study day 1. Participants were then excluded from the exploratory analysis if they were SARS-CoV-2 positive via salivary PCR at study day 1. Participants were further excluded from the Bayesian analysis where household size was greater than 7, as these household sizes were unable to be modelled using Bayesian modelling.

### Bayesian statistical analysis

Bayesian analysis was included in the original statistical analysis plan. The goal was to test the null hypothesis that treatment of household contacts with IFNβ-1α does not reduce the probability of transmission from an infected index case to household contacts. As outlined in the analysis plan, we used data to develop a data generating process governed by probabilities that a household contact of an index case would test positive to COVID-19, and non-informative prior was used.

### Bayesian model

A generalized linear model with mixed effects was developed to estimate the probability of infection for each contact. A stan_glmer with binomial logit function from the rstanarm package (1) was run using R (version 4.1.1) to obtain Beta coefficients for 4,000 simulated samples to estimate the probability of infection using the following model:

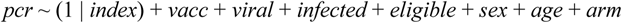

where: *pcr*: binary COVID-19 test result, *index*: index case in the household – treated as a random effect, *vacc*: number of vaccinated household members, *viral*: log_10_ viral load of the index case at the beginning of the period, *infected*: number of COVID-19 positive household members at the beginning of the period, *eligible*: whether the household contact was eligible for treatment, *sex*: sex of the household contact, *age*: age of the household contact, and *arm*: study arm (i.e. SOC/IFN) of the household were the fixed explanatory variables.

The features of the model were the same as the exploratory frequentist analysis, with the exception of adding the treatment period as a feature. Instead, the Bayesian analysis split the data into two periods (Period 1: study days 1–11, Period 2: study days 12–29) with analysis conducted separately. For period 2, COVID-19 positivity at day 11 was used to determine COVID-19 positive household members at the beginning of the period. The population used for analysis was the same as the “At-risk” population used in the exploratory frequentist analysis (Supplementary Table 6), with the additional exclusion of participants in households with a size larger than 8.

### Computation of difference in probability of infection in IFN vs SOC arms

In each simulation, we obtained a PCR outcome for each contact from the Bayesian model by applying the posterior_predict function in the rstanarm package to the observed data (1). We further computed the percent of infections in each treatment arm irrespective of household size (Supplementary Table 8) and for each household size (Supplementary Table 9). Furthermore, we computed the difference in probability of infection by subtracting the probability of infection in IFN arm from the probability of infection in SOC arm. As a results, we obtained 4000 differences (irrespective of household size) and 4000 differences for each household size

Significance of the treatment effect was determined by 95% credible interval (CrI) for the treatment Beta coefficient. If 0 was included in the CrI, the effect of treatment group was considered not significant. Average Beta coefficient and odds ratio for the effect of the IFN treatment as compared to SOC were computed in each study period. The average odds ratio of being infected with SARS-CoV-2 on IFN treatment vs. SOC was calculated by exponentiating the values of the average of the coefficients (Supplementary Table 7).

## Supplementary Results

### Bayesian analysis of infection reduction

Treatment with IFN had a significant effect in study period 1 as the 95% CrI for treatment coefficient did not include zero (Supplementary Table 7). However, IFN treatment did not have a significant effect in period 2 as the 95% CrI did include zero (Supplementary Table 7).

In study period 1, there was 95% probability of infection reduction between 0.9% to 15.9% due to the IFN treatment, with 8.5% median and mean infection reduction (Supplementary Table 8). In study period 2, there are 95% probability of infection reduction −6.7% to 4.8% due to the IFN treatment, with −9% of median and mean of infection reduction (Supplementary Table 8).

When we stratified the infection reduction by household size, only households’ size 3 and 4 in period 1 had a 95% CrI without zero (Supplementary Table 9).

As the distribution of Bayesian estimated differences in probabilities of infection in study period 1 is centred to the right of 0 (Supplementary Figure 3A), IFN treatment had an effect on reducing SARS-CoV-2 transmission within households. However, as the distribution in study period 2 is centred around 0, it suggests IFN has no effect on SARS-CoV-2 transmission within households (Supplementary Figure 3B).

## Supplementary Figures

**Supplementary Figure 1:**
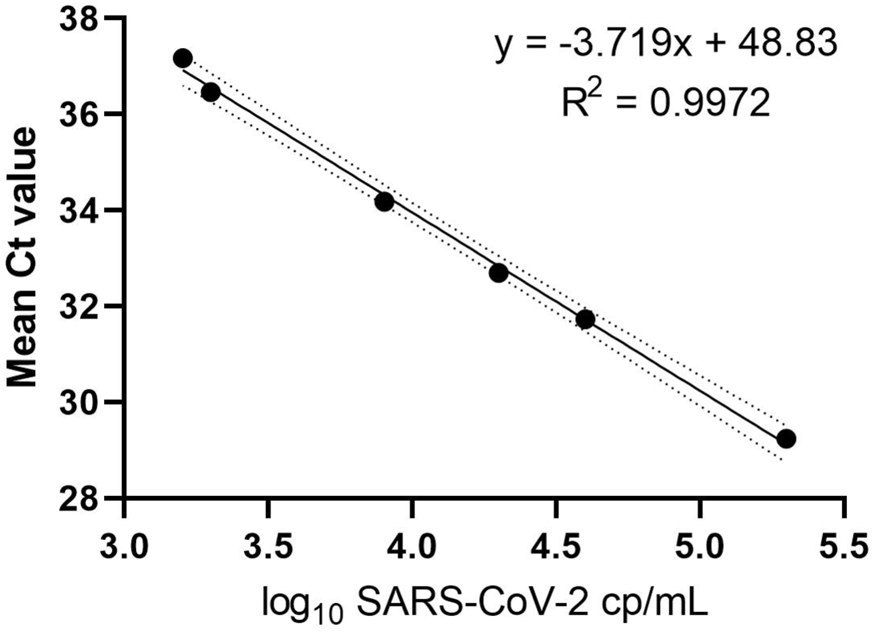
Standard curve for calculation of SARS-CoV-2 viral load. Mean cycle threshold (Ct) values are plotted on the y-axis, and log_10_ SARS-CoV-2 copies per mL (cp/mL) are plotted on the x-axis. Linear regression resulted in an R^2^ value of 0.9972, to fit the equation y = –3.719x + 48.83.

**Supplementary Figure 2:**
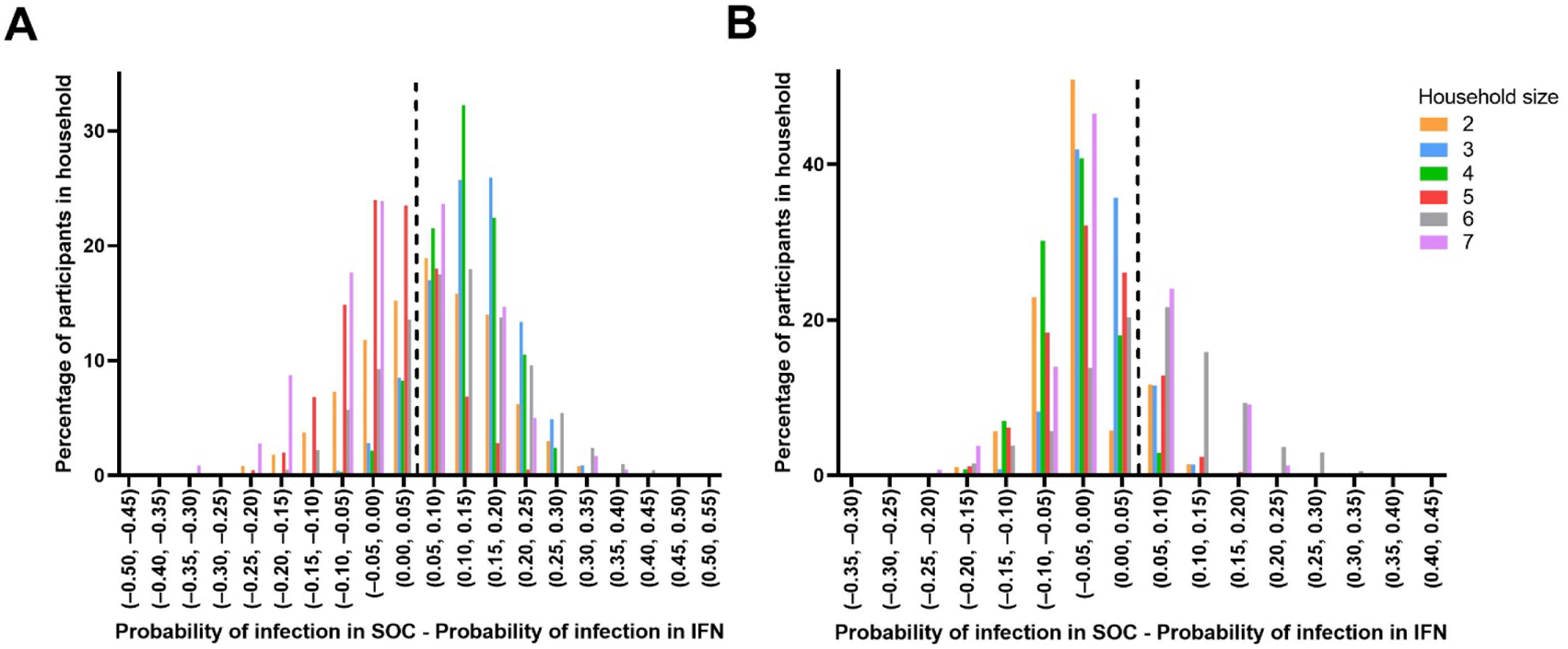
Distribution of the difference in probability of infection between SOC and IFN arms by household size. **(A)** Distribution in study period 1, and **(B)** Distribution in study period 2. Vertical dashed black line represents zero difference in probability of infection between SOC and IFN arms.

**Supplementary Figure 3:**
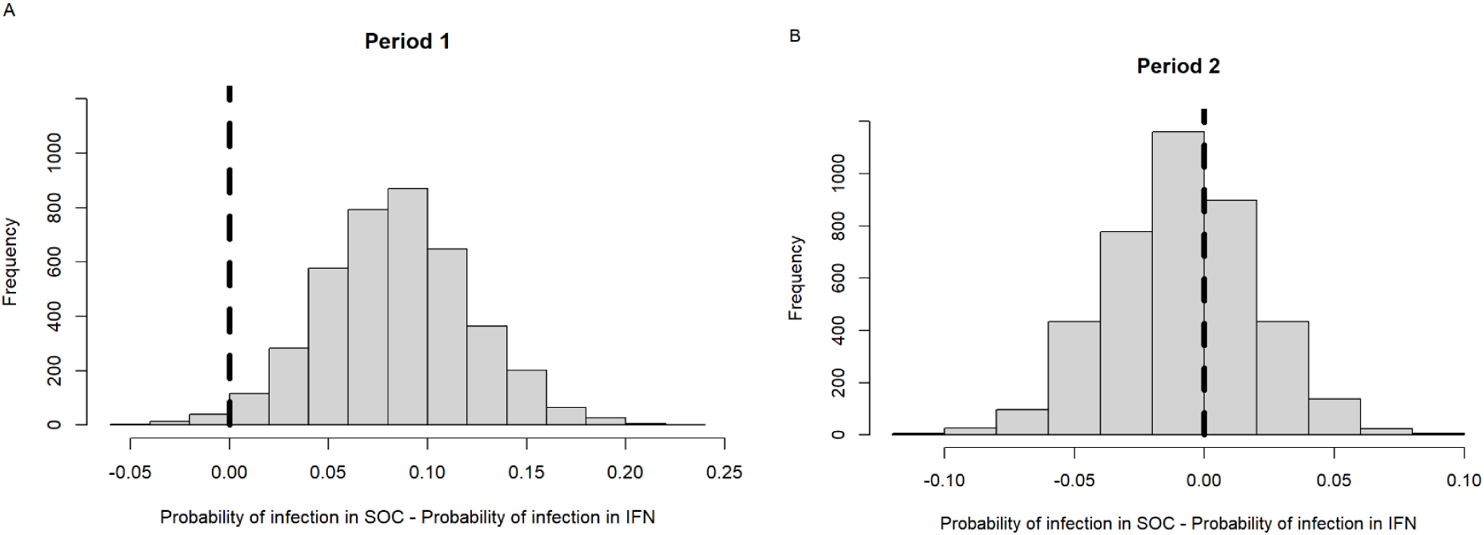
Distribution of the difference in probability of infection between SOC and IFN arms. (A) Distribution in study period 1, and (B) Distribution in study period 2. Vertical black line represents zero difference in probability of infection between SOC and IFN arms.

**Supplementary Figure 4:**
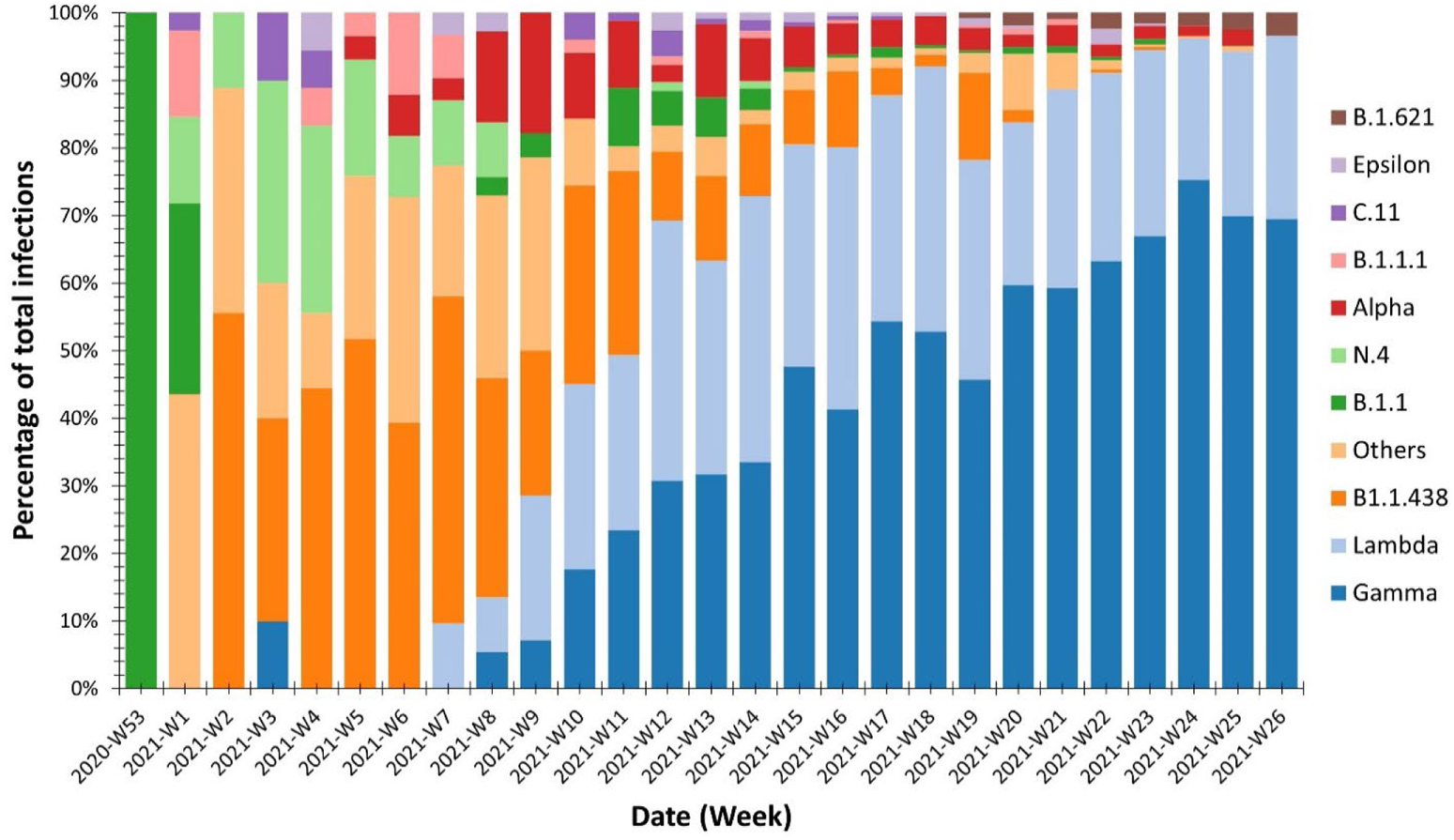
COVID-19 variants detected in Santiago, Chile during study recruitment. Data represents COVID-19 variants detected as a percentage of the total infections detected during this period.

## Supplementary Tables

**Supplementary Table 1:**
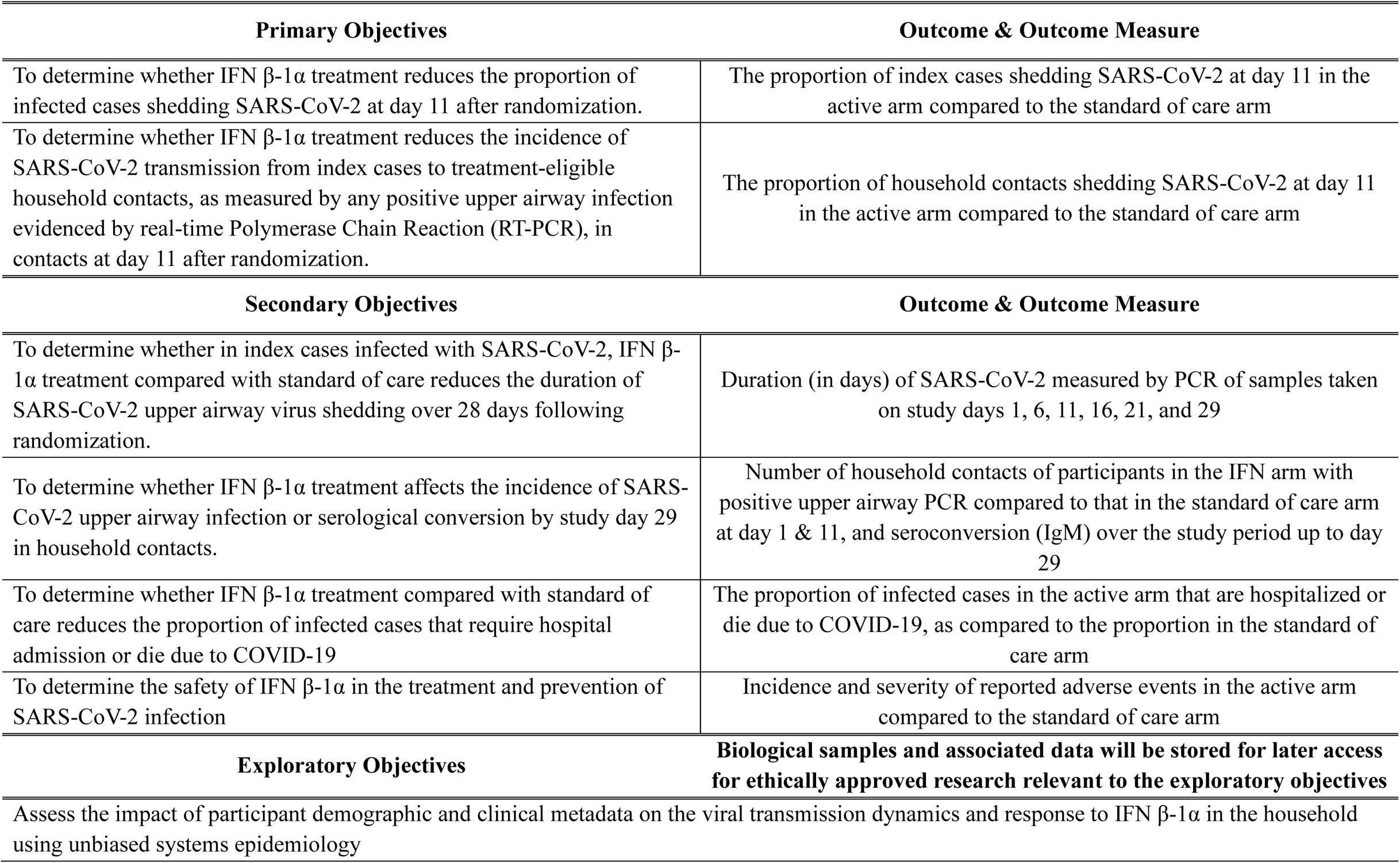
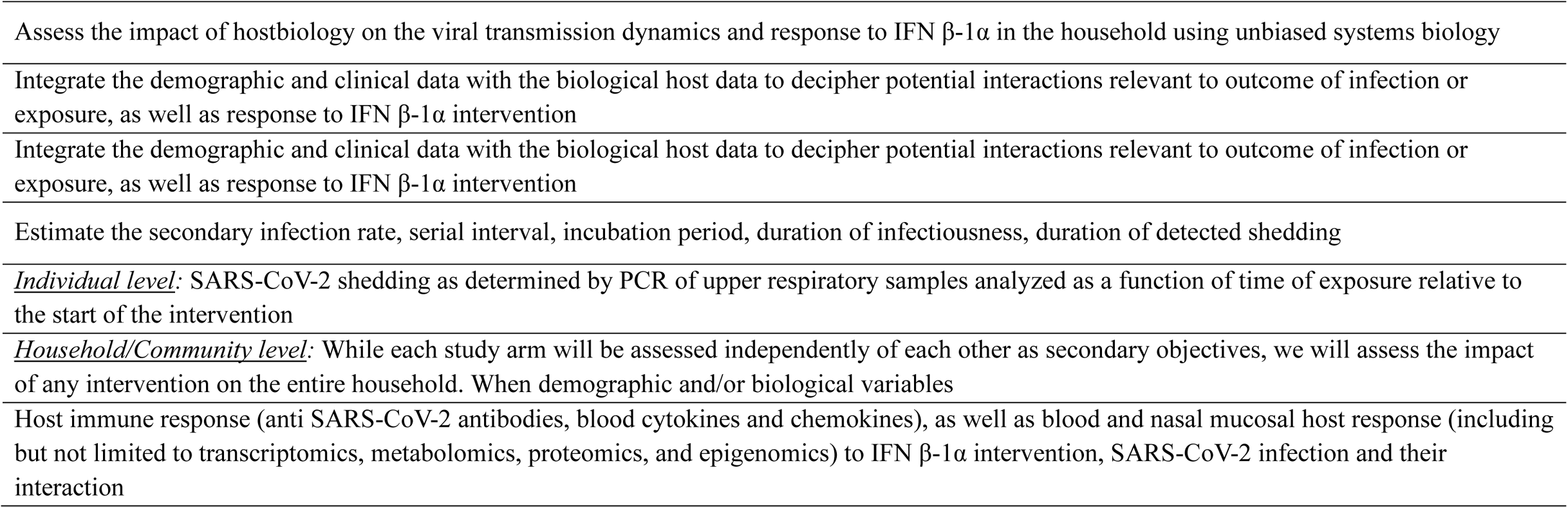
Primary, secondary, and exploratory outcomes for the Containing Coronavirus Disease-19 clinical trial.

| Primary Objectives | Outcome & Outcome Measure |
| --- | --- |
| To determine whether IFN β-1α treatment reduces the proportion of infected cases shedding SARS-CoV-2 at day 11 after randomization. | The proportion of index cases shedding SARS-CoV-2 at day 11 in the active arm compared to the standard of care arm |
| To determine whether IFN β-1α treatment reduces the incidence of SARS-CoV-2 transmission from index cases to treatment-eligible household contacts, as measured by any positive upper airway infection evidenced by real-time Polymerase Chain Reaction (RT-PCR), in contacts at day 11 after randomization. | The proportion of household contacts shedding SARS-CoV-2 at day 11 in the active arm compared to the standard of care arm |
| Secondary Objectives | Outcome & Outcome Measure |
| To determine whether in index cases infected with SARS-CoV-2, IFN β-1α treatment compared with standard of care reduces the duration of SARS-CoV-2 upper airway virus shedding over 28 days following randomization. | Duration (in days) of SARS-CoV-2 measured by PCR of samples taken on study days 1, 6, 11, 16, 21, and 29 |
| To determine whether IFN β-1α treatment affects the incidence of SARS-CoV-2 upper airway infection or serological conversion by study day 29 in household contacts. | Number of household contacts of participants in the IFN arm with positive upper airway PCR compared to that in the standard of care arm at day 1 & 11, and seroconversion (IgM) over the study period up to day 29 |
| To determine whether IFN β-1α treatment compared with standard of care reduces the proportion of infected cases that require hospital admission or die due to COVID-19 | The proportion of infected cases in the active arm that are hospitalized or die due to COVID-19, as compared to the proportion in the standard of care arm |
| To determine the safety of IFN β-1α in the treatment and prevention of SARS-CoV-2 infection | Incidence and severity of reported adverse events in the active arm compared to the standard of care arm |
| Exploratory Objectives | Biological samples and associated data will be stored for later access for ethically approved research relevant to the exploratory objectives |
| Assess the impact of participant demographic and clinical metadata on the viral transmission dynamics and response to IFN β-1α in the household using unbiased systems epidemiology | |

|  |
| --- |
| Assess the impact of hostbiology on the viral transmission dynamics and response to IFN β-1α in the household using unbiased systems biology |
| Integrate the demographic and clinical data with the biological host data to decipher potential interactions relevant to outcome of infection or exposure, as well as response to IFN β-1α intervention |
| Integrate the demographic and clinical data with the biological host data to decipher potential interactions relevant to outcome of infection or exposure, as well as response to IFN β-1α intervention |
| Estimate the secondary infection rate, serial interval, incubation period, duration of infectiousness, duration of detected shedding |
| *Individual level:* SARS-CoV-2 shedding as determined by PCR of upper respiratory samples analyzed as a function of time of exposure relative to the start of the intervention |
| *Household/Community level:* While each study arm will be assessed independently of each other as secondary objectives, we will assess the impact of any intervention on the entire household. When demographic and/or biological variables |
| Host immune response (anti SARS-CoV-2 antibodies, blood cytokines and chemokines), as well as blood and nasal mucosal host response (including but not limited to transcriptomics, metabolomics, proteomics, and epigenomics) to IFN β-1α intervention, SARS-CoV-2 infection and their interaction |
| **Supplementary Table 1: Primary, secondary, and exploratory outcomes for the Containing Coronavirus Disease-19 clinical trial** |

**Supplementary Table 2:** Schedule of laboratory assessments for index cases and treatment eligible household contacts.

|  | Visit 1 | Visit 2 | Visit 3 | Visit 4 | Visit 5 | Visit 6 | Extra Visit<br>– Suspected<br>COVID-19 |
| --- | --- | --- | --- | --- | --- | --- | --- |
| Study Day | 1<br>Dose 1 | 6<br>Dose 2 | 11<br>Dose 3 | 16 | 21 | 29 |  |
| **Blood Samples** |  |  |  |  |  |  |  |
| Complete Blood count/ ESR | X |  |  |  |  | X | X |
| SARS-CoV-2 antibodies (IgGAM) | X |  |  |  |  | X |  |
| Study sample 1 | X | X | X |  |  | X | X |
| Study sample 2 | X | X | X |  |  | X | X |
| Study sample 3 | X | X | X |  |  | X | X |
| **Airway samples** |  |  |  |  |  |  |  |
| Nasal brush | X |  |  |  |  |  |  |
| Nasal lining fluid | X | X | X |  |  |  |  |
| Airway Swab for COVID-19 diagnosis |  |  |  |  |  |  | X |
| **Saliva Samples** |  |  |  |  |  |  |  |
| Saliva | X | X | X | X | X | X | X |

**Supplementary Table 3:** Schedule of laboratory assessments for treatment ineligible household contacts.

|  | Visit 1 | Visit 2 | Visit 3 | Visit 4 | Visit 5 | Visit 6 | Extra Visit<br>– Suspected<br>COVID-19 |
| --- | --- | --- | --- | --- | --- | --- | --- |
| Study Day | 1<br>Dose 1 | 6<br>Dose 2 | 11<br>Dose 3 | 16 | 21 | 29 |  |
| **Blood Samples** |  |  |  |  |  |  |  |
| Complete Blood count/ ESR |  |  |  |  |  |  | X |
| SARS-CoV-2 antibodies (IgGAM) |  |  |  |  |  | X |  |
| Study sample 1 |  |  |  |  |  | X | X |
| Study sample 2 |  |  |  |  |  | X | X |
| Study sample 3 |  |  |  |  |  | X | X |
| **Airway samples** |  |  |  |  |  |  |  |
| Nasal brush | X |  |  |  |  |  |  |
| Nasal lining fluid | X | X | X |  |  |  |  |
| Airway Swab for COVID-19 diagnosis |  |  |  |  |  |  | X |
| **Saliva Samples** |  |  |  |  |  |  |  |
| Saliva | X | X | X | X | X | X | X |

**Supplementary Table 4:** Study withdrawal reasons by treatment arm.

| Interferon arm |  |  |  |  |  |  |  |  |  | Standard of Care arm |  |  |  |  |  |  |  |  |  |
| --- | --- | --- | --- | --- | --- | --- | --- | --- | --- | --- | --- | --- | --- | --- | --- | --- | --- | --- | --- |
| Withdrawal reason | Days since enrolment |  |  |  |  |  |  |  |  | Withdrawal reason | Days since enrolment |  |  |  |  |  |  |  |  |
|  | 3 | 4 | 5 | 10 | 11 | 14 | 15 | 21 | 28 |  | 3 | 4 | 5 | 10 | 11 | 14 | 15 | 25 | 28 |
| Family withdrew | 0 | 0 | 5 | 0 | 0 | 2 | 0 | 0 | 0 | Family withdrew | 0 | 0 | 2 | 0 | 0 | 0 | 0 | 0 | 0 |
| Interpersonal problems | 0 | 0 | 3 | 0 | 0 | 0 | 0 | 0 | 0 | Interpersonal problems | 0 | 4 | 0 | 0 | 0 | 0 | 0 | 0 | 0 |
| Lack of time | 0 | 1 | 0 | 0 | 0 | 0 | 0 | 0 | 0 | Lack of time | 0 | 0 | 0 | 0 | 0 | 0 | 0 | 0 | 0 |
| Other | 0 | 0 | 0 | 0 | 0 | 0 | 0 | 0 | 0 | Other | 0 | 0 | 1 | 0 | 0 | 0 | 0 | 0 | 0 |
| Parental decision | 0 | 0 | 2 | 0 | 0 | 0 | 0 | 0 | 0 | Parental decision | 0 | 0 | 0 | 0 | 0 | 0 | 0 | 0 | 0 |
| Poor health | 0 | 0 | 1 | 0 | 0 | 1 | 0 | 0 | 0 | Poor health | 0 | 0 | 0 | 0 | 0 | 0 | 0 | 0 | 0 |
| Sample collection problems | 0 | 0 | 0 | 0 | 0 | 0 | 0 | 0 | 0 | Sample collection problems | 0 | 0 | 1 | 0 | 0 | 0 | 0 | 0 | 0 |
| Side effects of drug | 0 | 0 | 0 | 0 | 0 | 2 | 0 | 0 | 0 | Side effects of drug | 0 | 0 | 0 | 0 | 0 | 0 | 0 | 0 | 0 |
| Uninterested | 0 | 0 | 0 | 1 | 0 | 0 | 0 | 0 | 1 | Uninterested | 0 | 0 | 0 | 0 | 0 | 0 | 0 | 0 | 0 |
| Unspecified | 4 | 3 | 1 | 2 | 1 | 0 | 0 | 0 | 0 | Unspecified | 0 | 1 | 3 | 0 | 0 | 0 | 1 | 5 | 0 |
| Work commitments | 0 | 0 | 2 | 2 | 0 | 0 | 0 | 0 | 0 | Work commitments | 0 | 0 | 0 | 0 | 0 | 0 | 0 | 0 | 0 |

**Supplementary Table 5:** Reasons for exclusion from study participation during screening and reasons for declined participation following invitation to participate in ConCorD-19.

| **Reasons for exclusion from ConCorD-19 during screening** | **Participants (n)** |
| --- | --- |
| Unable to be contacted | 736 |
| Days of symptoms > 5 | 777 |
| Asymptomatic | 306 |
| Isolating from household contacts | 219 |
| Hospitalized | 177 |
| Confirmation of COVID-19 took longer than 72 hours | 35 |
| Deceased | 2 |
| Ineligible medical condition or treatment | 67 |
| Less than 18 years old | 54 |
| Pregnant, or plans on becoming pregnant | 32 |
| Previously infected with COVID-19 | 36 |
| Greater than 80 years old | 3 |
| All eligible household contacts currently infected | 514 |
| Does not live with eligible household contact | 412 |
| Not the first infection in the household | 392 |
| Does not live in the Santiago metropolitan area | 20 |
| **Reasons for not participating in ConCorD-19** |  |
| Index case declined | 835 |
| Eligible household contact declined | 196 |
| **Total** | **4813** |

**Supplementary Table 6:**
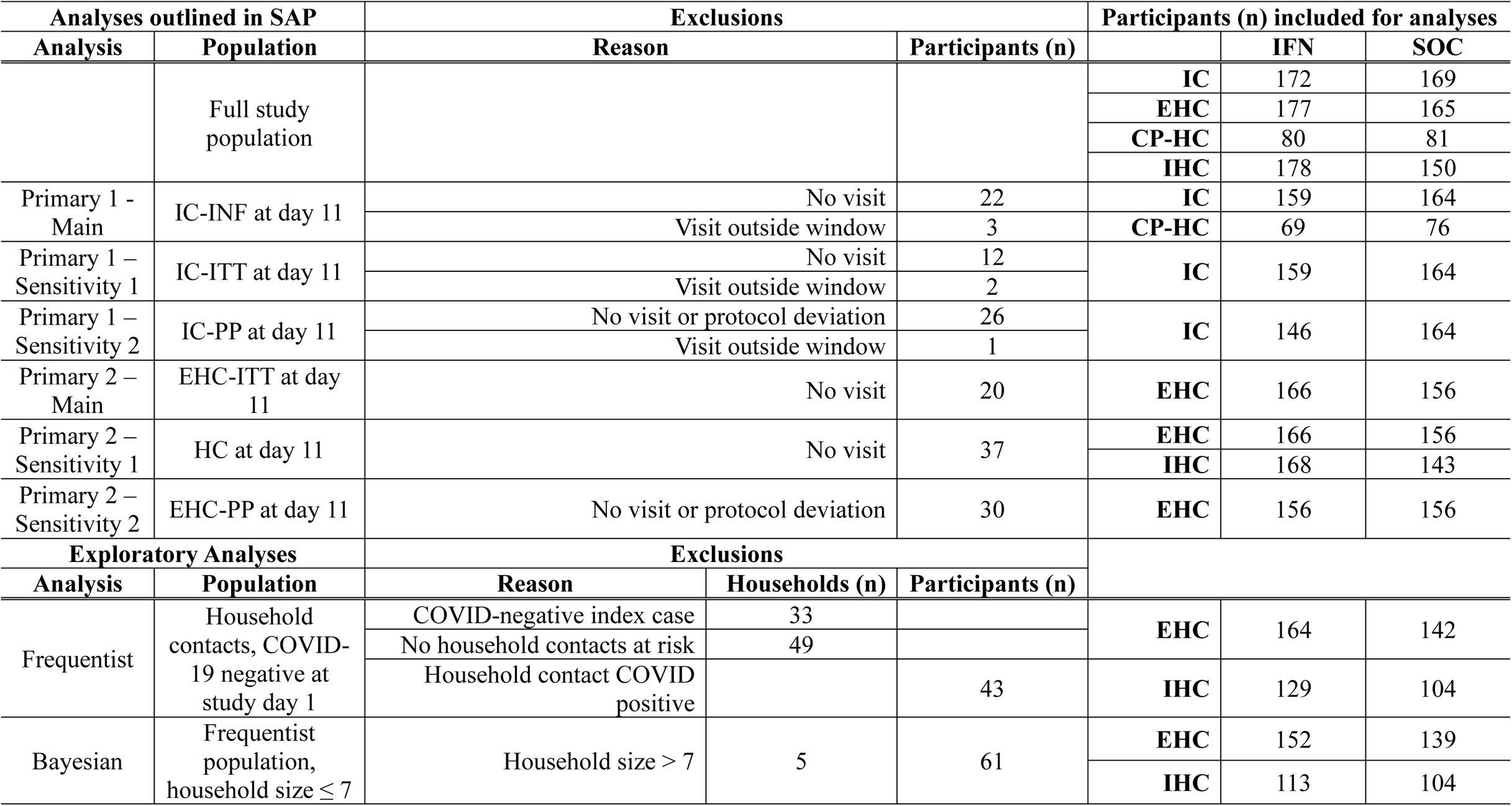
Study populations used for analysis, and reasons for exclusion from analysis. IC: index case, EHC: Treatment eligible household contacts, CP-HC: COVID-19 positive household contacts, IHC: Treatment ineligible household contacts, IC-INF: IC & CP-HC, IC-ITT: Index case – intention to treat population, IC-PP: Index case – per protocol population, EHC-ITT: Treatment eligible household contacts – intention to treat population, HC: EHC and IHC, EHC-PP: Treatment eligible household contact – per protocol population.

**Supplementary Table 7:** Bayesian estimated beta coefficients and odds ratio for treatment with IFN, with average values and 95% credible intervals.

| Coefficient for treatment with IFN |  |  | 95% Credible Intervals |  |
| --- | --- | --- | --- | --- |
| Study period | Mean | Standard Deviation | Lower limit (2.5%) | Upper limit (97.5%) |
| 1 | -1.17 | 0.52 | -2.23 | -0.19 |
| 2 | 0.27 | 0.55 | -0.80 | 1.36 |

| Odds ratio for treatment with IFN |  |  | 95% Credible Intervals |  |
| --- | --- | --- | --- | --- |
| Study period | Mean | Standard Deviation | Lower limit (2.5%) | Upper limit (97.5%) |
| 1 | 0.32 | 1.68 | 0.11 | 0.83 |
| 2 | 1.31 | 1.73 | 0.45 | 3.90 |

**Supplementary Table 8:** Difference in probability of infection in each study period irrespective of household size. Data is the mean, median, and 95% CrI difference in probability of infection in each study period, computed as probability of infection in the SOC arm minus the probability of infection in the IFN arm.

| Study period | 2.5% | 25% | 50% | Mean | 75% | 97.5% |
| --- | --- | --- | --- | --- | --- | --- |
| 1 | 0.009 | 0.059 | 0.085 | 0.085 | 0.110 | 0.159 |
| 2 | -0.067 | -0.028 | -0.009 | -0.009 | 0.010 | 0.048 |

**Supplementary Table 9:** Difference in probability of infection in each study period, stratifying by household size. Data is the mean, median, and 95% CrI difference in probability of infection in each study period, computed as probability of infection in the SOC arm minus the probability of infection in the IFN arm.

| Study period | Household size | 2.5% | 25% | 50% | Mean | 75% | 97.5% |
| --- | --- | --- | --- | --- | --- | --- | --- |
| 1 | 2 | -0.143 | 0.000 | 0.286 | 0.053 | 0.143 | 0.286 |
| 1 | 3 | 0.000 | 0.092 | 0.276 | 0.139 | 0.184 | 0.276 |
| 1 | 4 | 0.003 | 0.087 | 0.249 | 0.128 | 0.170 | 0.249 |
| 1 | 5 | -0.146 | -0.046 | 0.158 | 0.005 | 0.058 | 0.158 |
| 1 | 6 | -0.120 | 0.030 | 0.326 | 0.103 | 0.189 | 0.326 |
| 1 | 7 | -0.231 | -0.077 | 0.308 | 0.024 | 0.077 | 0.308 |
| 2 | 2 | -0.118 | -0.059 | 0.075 | -0.008 | 0.000 | 0.075 |
| 2 | 3 | -0.085 | -0.021 | 0.090 | 0.001 | 0.028 | 0.090 |
| 2 | 4 | -0.130 | -0.068 | 0.059 | -0.037 | -0.006 | 0.059 |
| 2 | 5 | -0.128 | -0.052 | 0.103 | -0.013 | 0.027 | 0.103 |
| 2 | 6 | -0.129 | -0.015 | 0.254 | 0.059 | 0.117 | 0.254 |
| 2 | 7 | -0.154 | 0.000 | 0.154 | 0.016 | 0.077 | 0.154 |

**Supplementary Table 10:** Number of contacts at each household size.

| Household size | Number of households | Eligible Contacts | Ineligible Contacts |
| --- | --- | --- | --- |
| 2 | 42 | 42 | 0 |
| 3 | 61 | 64 | 27 |
| 4 | 80 | 84 | 89 |
| 5 | 46 | 58 | 59 |
| 6 | 19 | 29 | 30 |
| 7 | 6 | 14 | 12 |

## Supplementary Appendices

### Supplementary Appendices A

**Containing Coronavirus Disease 19 Trial (CONCORD-19): Statistical Analysis Report**

Statistical Analysis Report

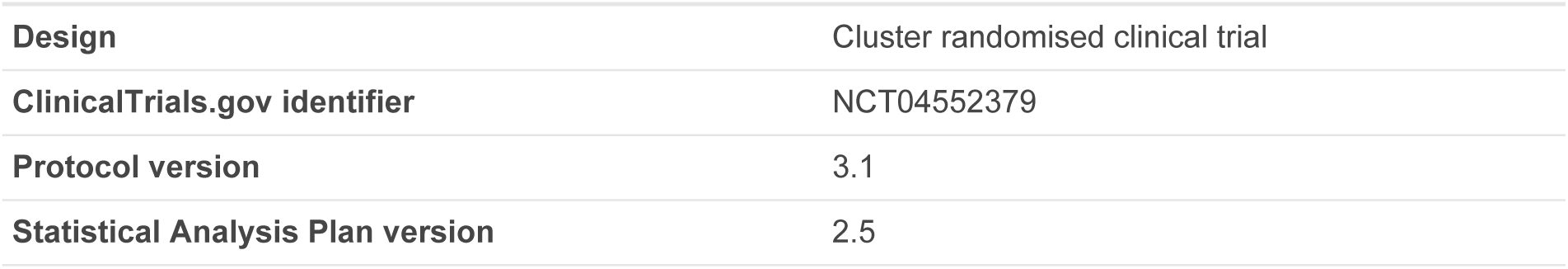

|  |  |
| --- | --- |
| **Design** | Cluster randomised clinical trial |
| **ClinicalTrials.gov identifier** | NCT04552379 |
| **Protocol version** | 3.1 |
| **Statistical Analysis Plan version** | 2.5 |

**Principal Investigators**

Stephen Stick, MB BChir PhD MRCP FRACP

Perth Children’s Hospital

15 Hospital Avenue, Nedlands WA 6009, Australia

Jose Antonio Castro-Rodriguez, MD PhD

School of Medicine, Pontificia Universidad Católica de Chile

Diagonal Paraguay 367, Santiago, Chile

**Author**

Virginia Chen, MA

Senior Data Scientist

PROOF Centre of Excellence

Vancouver, BC, Canada

**Signatures**

**Statistician (Author)**

Virginia Chen

**Statistician (Reviewer)**

Robert Balshaw

**Statistician (Reviewer)**

Casey P. Shannon

## 1. Abbreviations

1 Abbreviations
|  |  |
| --- | --- |
| **AE** | adverse event |
| **AR** | absolute risk |
| **ARR** | absolute risk reduction |
| **GLM** | generalized linear model |
| **GLMM** | generalized linear mixed model |
| **HR** | hazard ratio |
| **IFN** | interferon beta 1alpha (Biogen) |
| **ITT** | intention-to-treat |
| **OR** | odds ratio |
| **PCR** | polymerase chain reaction |
| **RR** | risk ratio |
| **SAE** | serious adverse event |
| **SAP** | statistical analysis plan |
| **SARS-CoV-2** | severe acute respiratory syndrome coronavirus 2 |
| **SD** | standard deviation |
| **SOC** | standard of care |
| **SOP** | standard operating procedure |

## 2. Introduction

### 2.1 Document Purpose

This document contains the results of the analyses described in the CONCORD-19 Statistical Analysis Plan (SAP). Descriptions of the analyses are omitted where the analysis proceeded as planned in the SAP. Any details or clarifying text are intended to be read in the context of the SAP.

### 2.2 Trial Objectives

#### 2.2.1 Primary Objectives

1. To determine whether IFN therapy reduces the number of infected cases still shedding SARS-CoV-2 in saliva at day 11 following randomisation.
2. To determine whether IFN therapy reduces the incidence of SARS CoV-2 transmission from index cases to treatment-eligible household contacts, as measured by any positive upper airway PCR in treatment-eligible household contacts at day 11 after randomisation

#### 2.2.2 Secondary Objectives

1. To determine whether in index cases infected with SARS-CoV-2, IFN therapy compared with SOC reduces the duration of a saliva positive polymerase chain reaction (PCR) SARS-CoV-2 test over 28 days following randomisation.
2. To determine if IFN affects the incidence of saliva positive PCR SARS-CoV-2 tests or serological conversion by study day 29 in household contacts, compared with SOC.
3. To determine if IFN therapy, compared with SOC, reduces the proportion of infected cases that require hospital admission or die due to COVID-19.
4. To determine the safety of IFN treatment.

#### 2.2.3 Exploratory Objectives

Exploratory objectives were not pursued at this stage.

### 2.3 Executive Summary

**Primary objective 1**

- Results do not suggest an effect

**Primary objective 2**

- Results suggest that IFN may reduce transmission to household contacts by study day 11, but statistical significance was not achieved in any analysis

**Secondary objective 1**

- Results do not suggest an effect

**Secondary objective 2**

- Results do not suggest an effect

**Secondary objective 3**

- Results suggest that IFN may increase duration of hospital stay, but statistical significance was not achieved in the main analysis
- IFN was statistically significant for reducing incidence of hospitalization in the sensitivity analyses
- IFN was statistically significant for increasing duration of hospital stay, in the sensitivity analyses

**Secondary objective 4**

- IFN was statistically significant for increasing incidence of treatment-related adverse events, but not for adverse events or serious adverse events in general

### 2.4 Deviations from the SAP

1. Two unplanned sensitivity analyses were added that take into account days between symptom onset and visit date for index cases. See “Adjusted for Days Between Symptom Onset and Visit Date” sections under “Sensitivity Analyses” for each Primary Objective.
2. Index cases occasionally had a negative PCR result on study day 1, despite testing positive prior to enrollment and testing positive at study day 6, 11, or 16. In these cases, their study day 1 viral load was imputed as 50% of their first non-negative viral load recorded during the study. These imputed values were used only in sensitivity analyses and additional analyses. Additionally, a sensitivity analysis will be performed for primary analysis 1, by excluding index cases that tested negative at *both* study day 1 and 6.
3. **An additional exploratory analysis into treatment effect during (D1-D11) and after (D12-29) discontinuation of treatment/household isolation is included (under Optional Analyses at the end of the report).**

### 2.5 Other Details

For inclusion as a covariate, age was discretised by quartile. The breakdown is as follows:

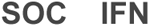
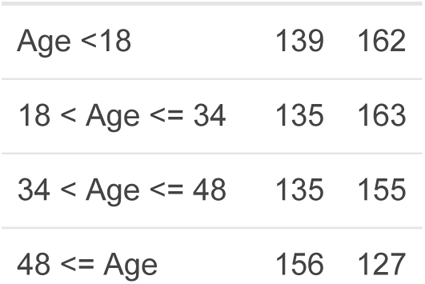

## 3 Analysis Populations and Participant Disposition

### 3.1 Full Analysis Population

3 Analysis Populations and Participant Disposition 3.1 Full Analysis Population
| IFN |  |  |  |  |  |  | SOC |  |  |  |  |  |  |
| --- | --- | --- | --- | --- | --- | --- | --- | --- | --- | --- | --- | --- | --- |
| # Visits conducted at study day |  |  |  |  |  |  | # Visits conducted at study day |  |  |  |  |  |  |
|  | 1 | 6 | 11 | 16 | 21 | 29 |  | 1 | 6 | 11 | 16 | 21 | 29 |
| Index case | 172 | 162 | 160 | 159 | 158 | 162 | Index case | 169 | 165 | 165 | 162 | 159 | 165 |
| Treatment-eligible household contact | 177 | 171 | 166 | 162 | 163 | 163 | Treatment-eligible household contact | 165 | 159 | 156 | 154 | 150 | 154 |
| COVID-positive household contact | 80 | 74 | 70 | 67 | 69 | 69 | COVID-positive household contact | 81 | 78 | 76 | 72 | 77 | 78 |
| Treatment-ineligible household contact | 178 | 173 | 168 | 169 | 169 | 167 | Treatment-ineligible household contact | 150 | 144 | 143 | 143 | 140 | 143 |

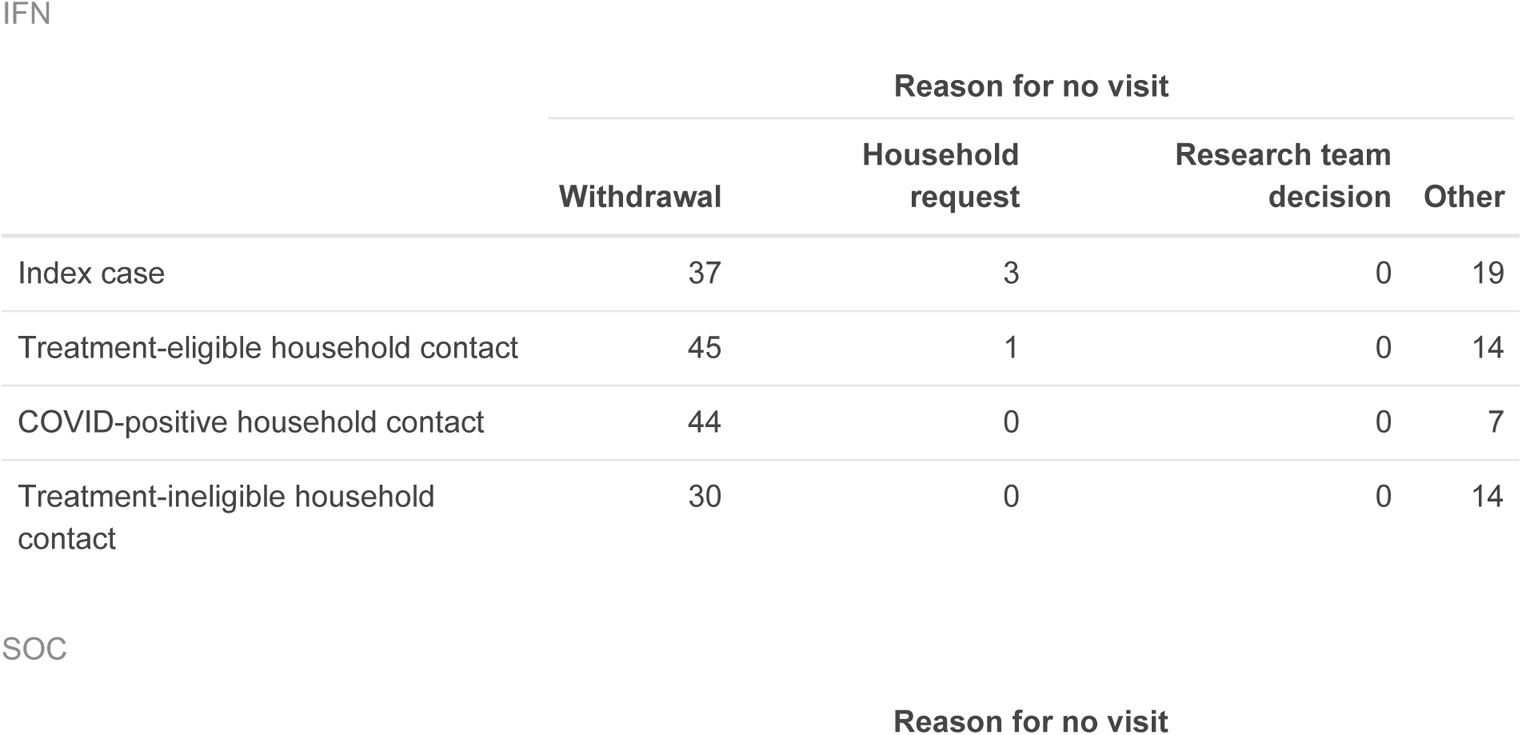
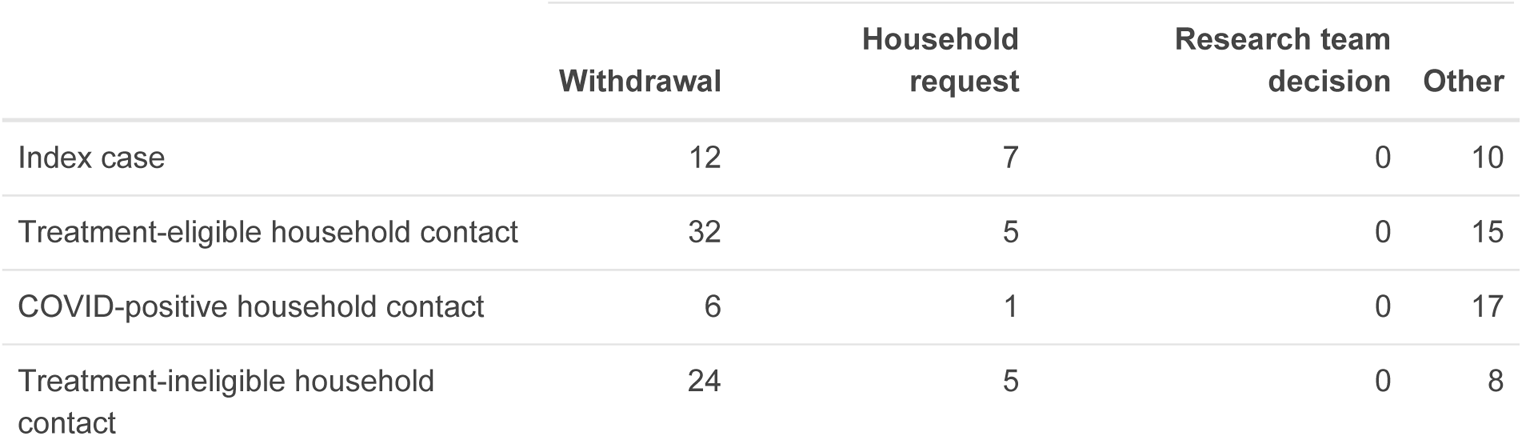

### 3.2 Index Case - Intention-to-treat Population IC-ITT

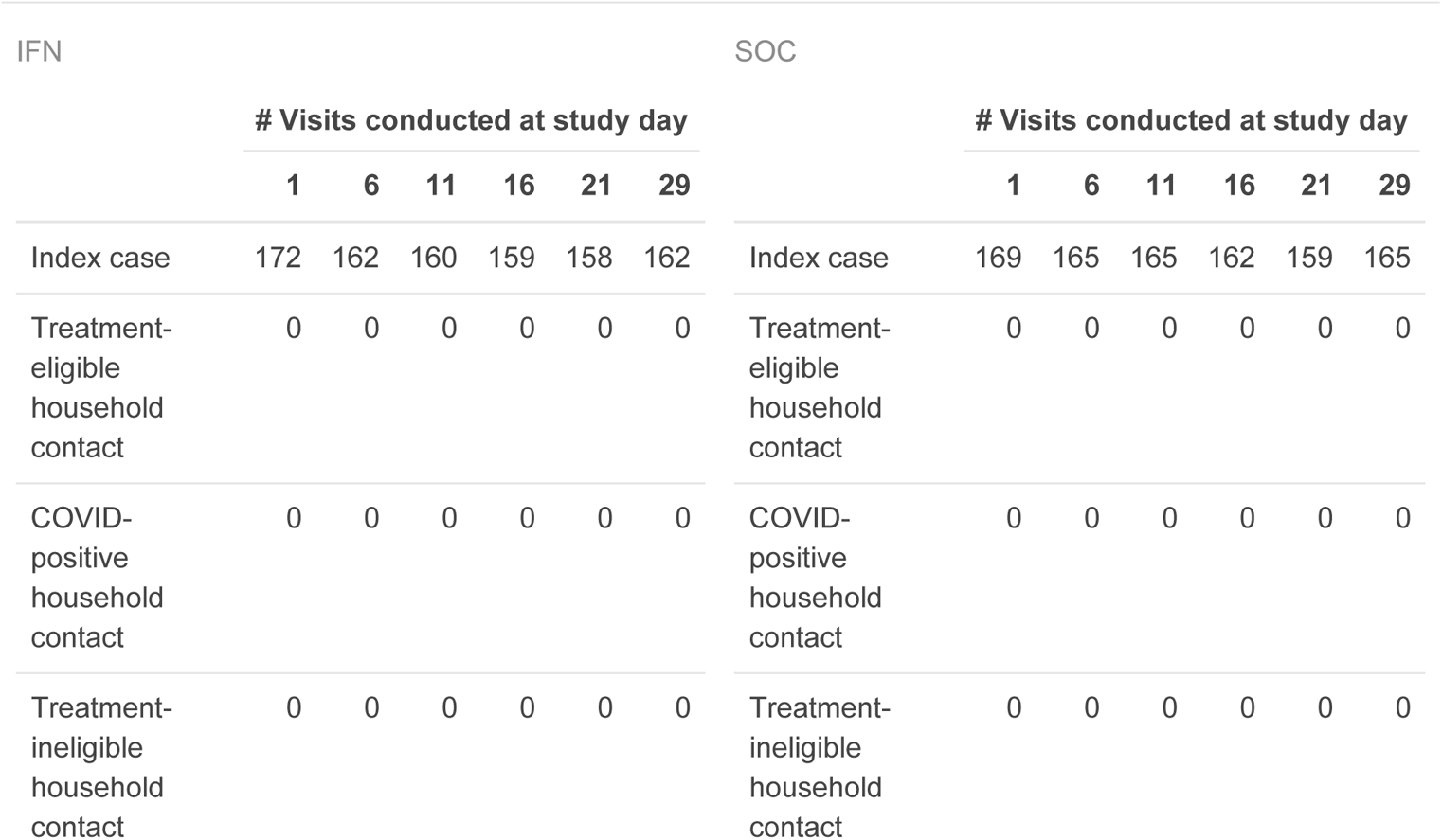
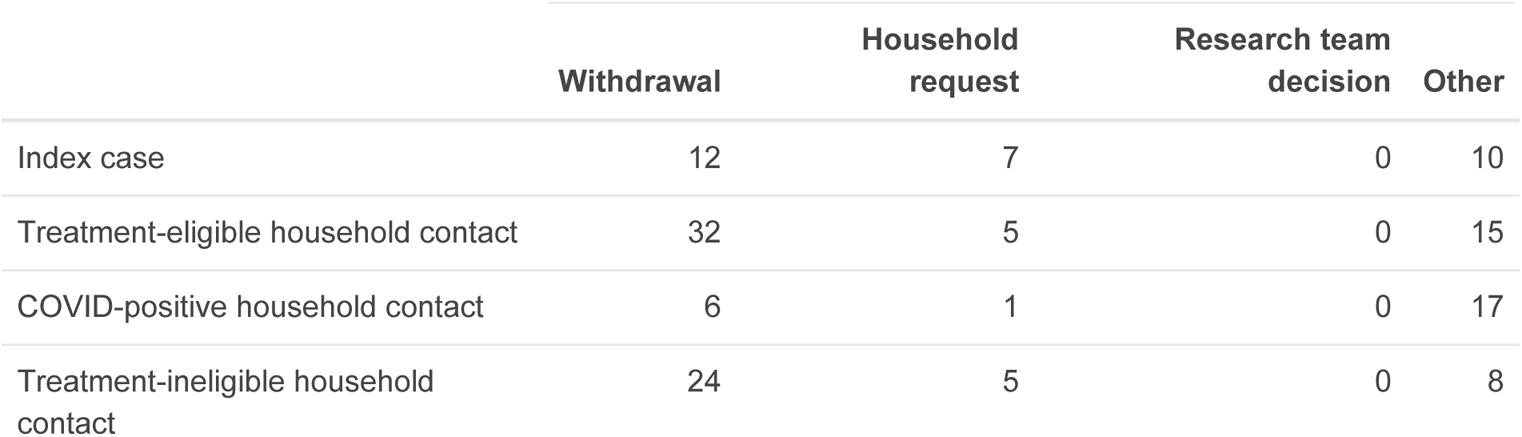

### 3.3 Index Case - Per Protocol Population IC-PP

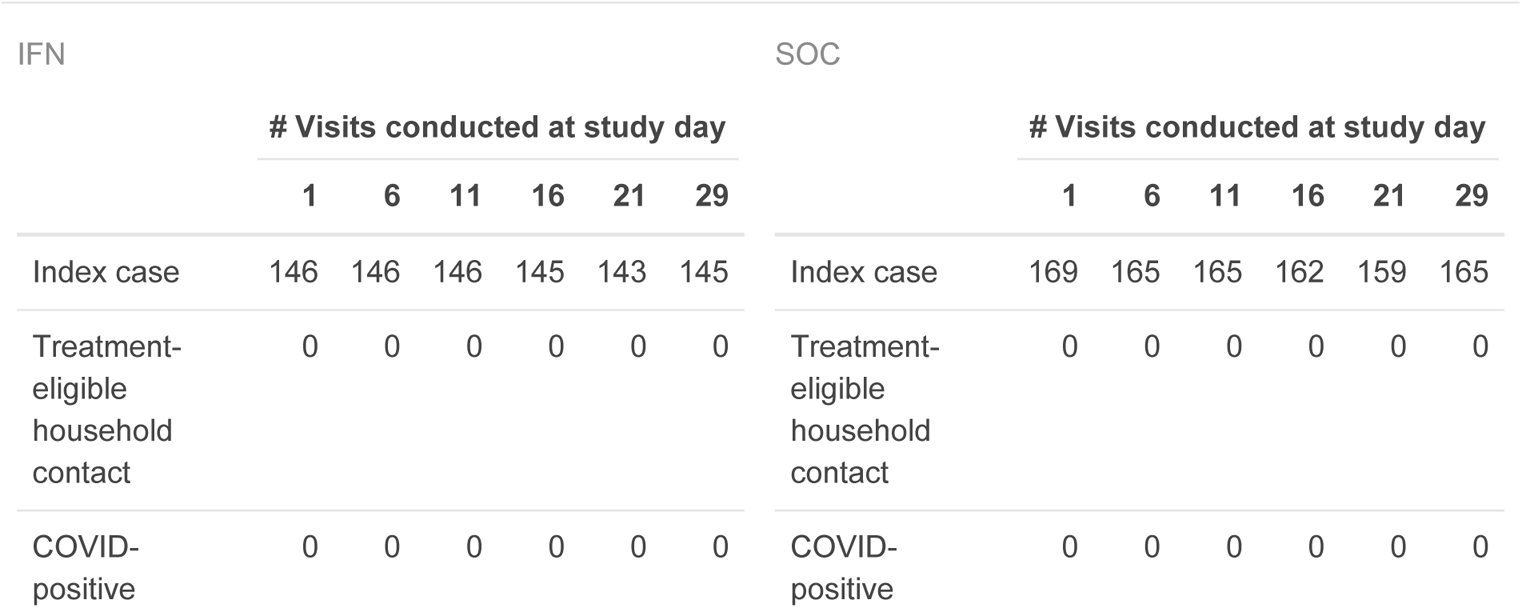
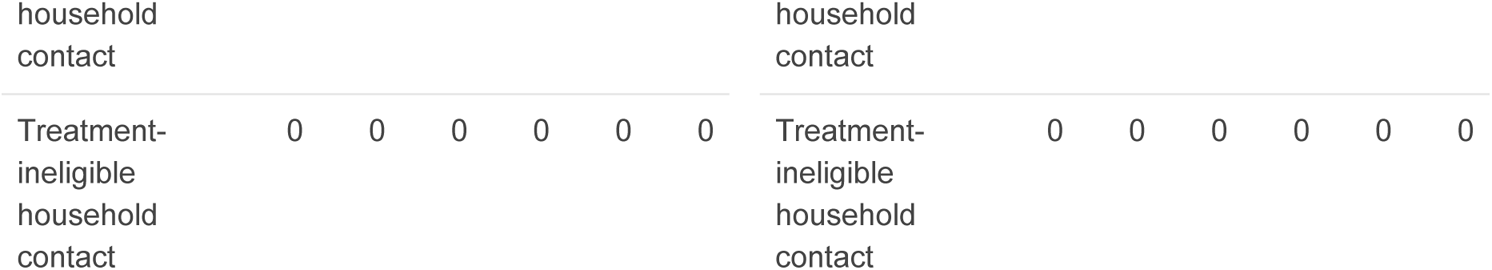

### 3.4 Infected Case Population IC-INF

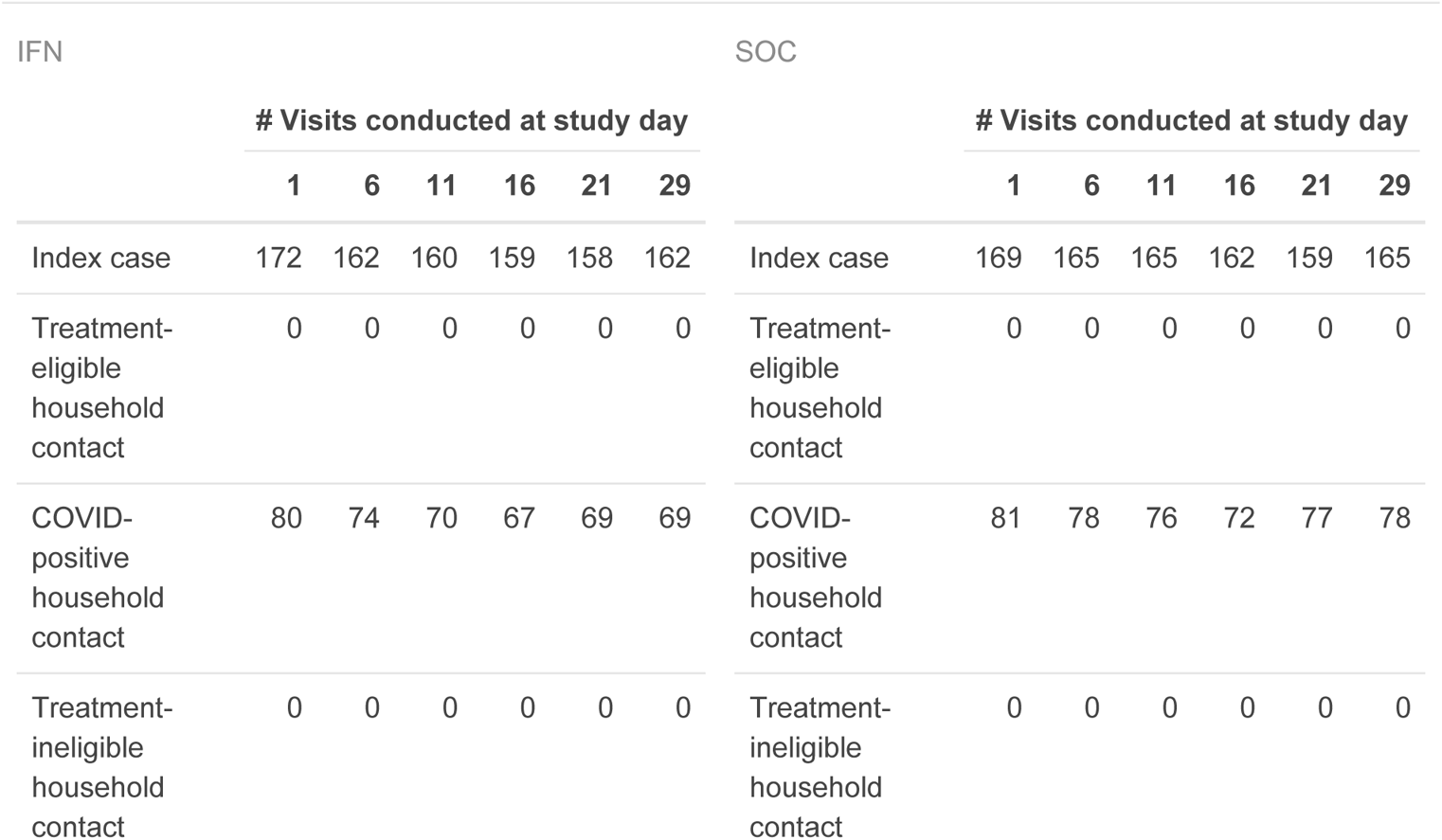

### 3.5 Treatment-eligible Household Contact - Intention-to-treat Population EHC-ITT

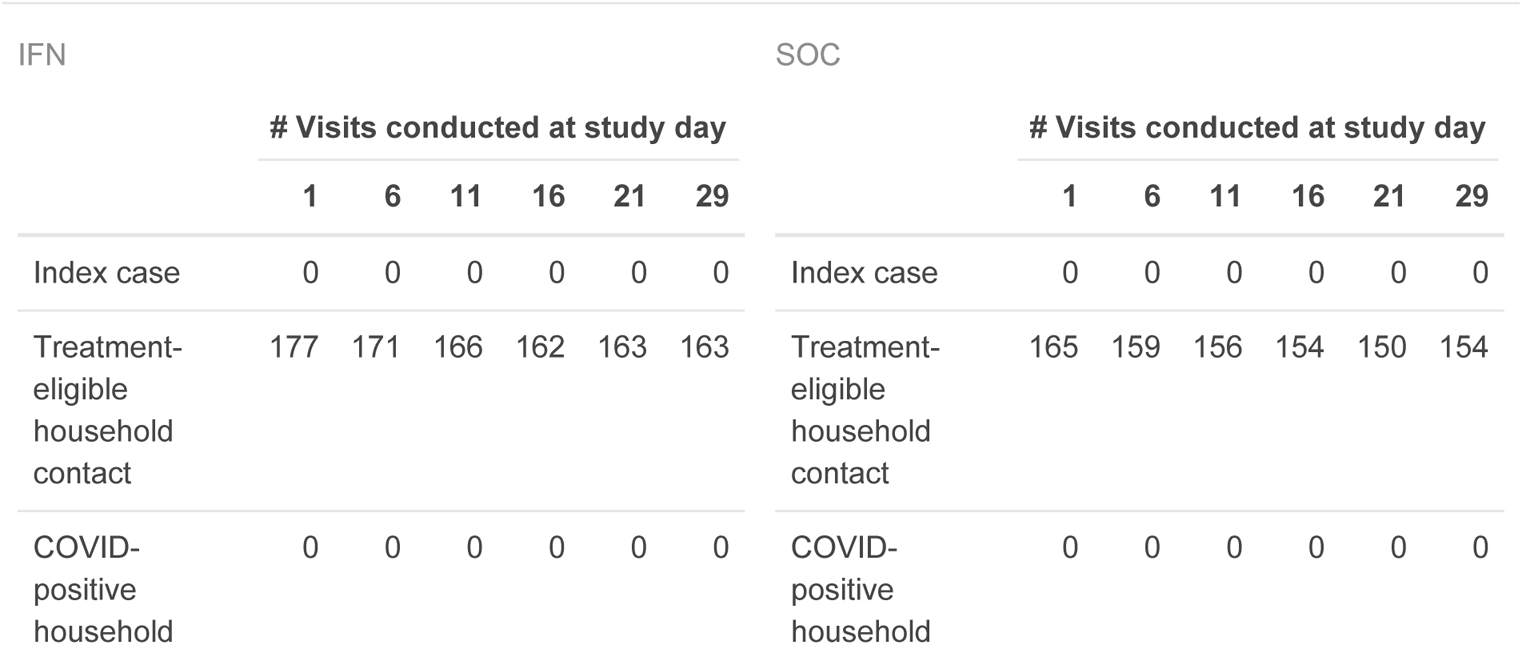
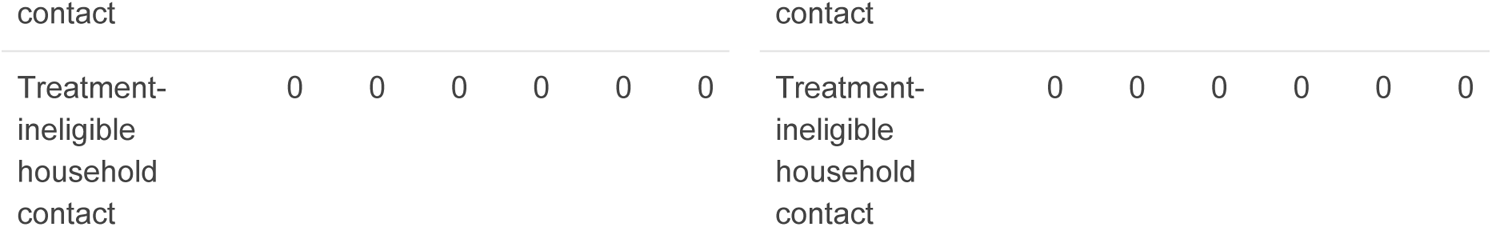

### 3.6 Treatment-eligible Household Contact - Per Protocol Population EHC-PP

| IFN |  |  |  |  |  |  | SOC |  |  |  |  |  |  |
| --- | --- | --- | --- | --- | --- | --- | --- | --- | --- | --- | --- | --- | --- |
| # Visits conducted at study day |  |  |  |  |  |  | # Visits conducted at study day |  |  |  |  |  |  |
|  | 1 | 6 | 11 | 16 | 21 | 29 |  | 1 | 6 | 11 | 16 | 21 | 29 |
| Index case | 0 | 0 | 0 | 0 | 0 | 0 | Index case | 0 | 0 | 0 | 0 | 0 | 0 |
| Treatment-eligible<br>household<br>contact | 156 | 156 | 156 | 152 | 154 | 153 | Treatment-eligible<br>household<br>contact | 165 | 159 | 156 | 154 | 150 | 154 |
| COVID-positive<br>household<br>contact | 0 | 0 | 0 | 0 | 0 | 0 | COVID-positive<br>household<br>contact | 0 | 0 | 0 | 0 | 0 | 0 |
| Treatment-ineligible<br>household<br>contact | 0 | 0 | 0 | 0 | 0 | 0 | Treatment-ineligible<br>household<br>contact | 0 | 0 | 0 | 0 | 0 | 0 |

### 3.7 Household Contact Population HC

| IFN |  |  |  |  |  |  | SOC |  |  |  |  |  |  |
| --- | --- | --- | --- | --- | --- | --- | --- | --- | --- | --- | --- | --- | --- |
| # Visits conducted at study day |  |  |  |  |  |  | # Visits conducted at study day |  |  |  |  |  |  |
|  | 1 | 6 | 11 | 16 | 21 | 29 |  | 1 | 6 | 11 | 16 | 21 | 29 |
| Index case | 0 | 0 | 0 | 0 | 0 | 0 | Index case | 0 | 0 | 0 | 0 | 0 | 0 |
| Treatment-eligible<br>household<br>contact | 177 | 171 | 166 | 162 | 163 | 163 | Treatment-eligible<br>household<br>contact | 165 | 159 | 156 | 154 | 150 | 154 |
| COVID-positive<br>household<br>contact | 0 | 0 | 0 | 0 | 0 | 0 | COVID-positive<br>household<br>contact | 0 | 0 | 0 | 0 | 0 | 0 |
| Treatment-ineligible<br>household<br>contact | 178 | 173 | 168 | 169 | 169 | 167 | Treatment-ineligible<br>household<br>contact | 150 | 144 | 143 | 143 | 140 | 143 |

### 3.8 Safety Population

| IFN |  |  |  |  |  |  | SOC |  |  |  |  |  |  |
| --- | --- | --- | --- | --- | --- | --- | --- | --- | --- | --- | --- | --- | --- |
|  | # Visits conducted at study day |  |  |  |  |  |  | # Visits conducted at study day |  |  |  |  |  |
|  | 1 | 6 | 11 | 16 | 21 | 29 |  | 1 | 6 | 11 | 16 | 21 | 29 |
| Index case | 172 | 162 | 160 | 159 | 158 | 162 | Index case | 169 | 165 | 165 | 162 | 159 | 165 |
| Treatment-eligible<br>household<br>contact | 177 | 171 | 166 | 162 | 163 | 163 | Treatment-eligible<br>household<br>contact | 165 | 159 | 156 | 154 | 150 | 154 |
| COVID-positive<br>household<br>contact | 80 | 74 | 70 | 67 | 69 | 69 | COVID-positive<br>household<br>contact | 81 | 78 | 76 | 72 | 77 | 78 |
| Treatment-ineligible<br>household<br>contact | 0 | 0 | 0 | 0 | 0 | 0 | Treatment-ineligible<br>household<br>contact | 0 | 0 | 0 | 0 | 0 | 0 |

## 4 Withdrawals

### 4.1 Overall

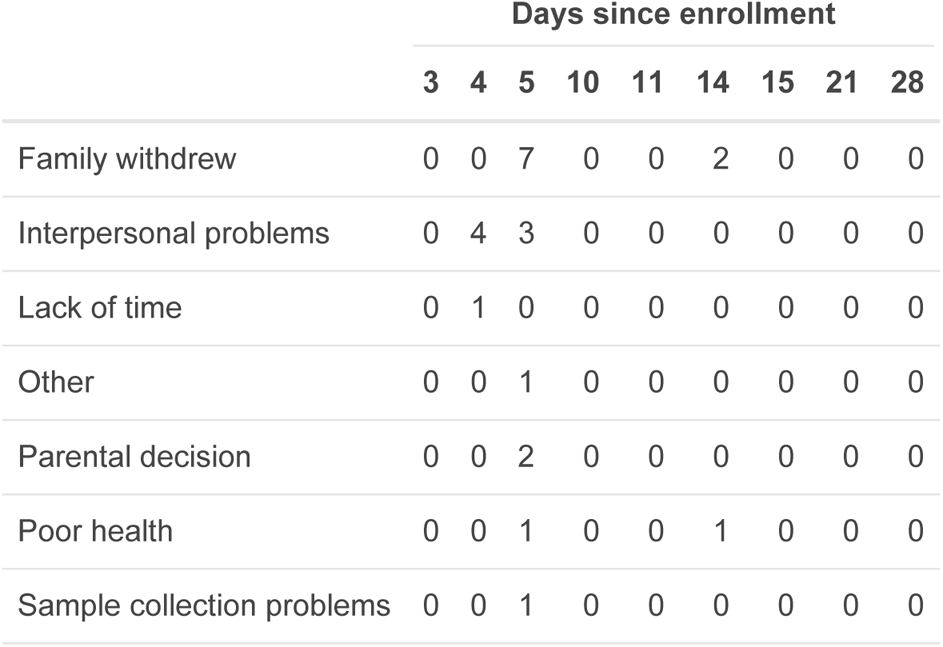
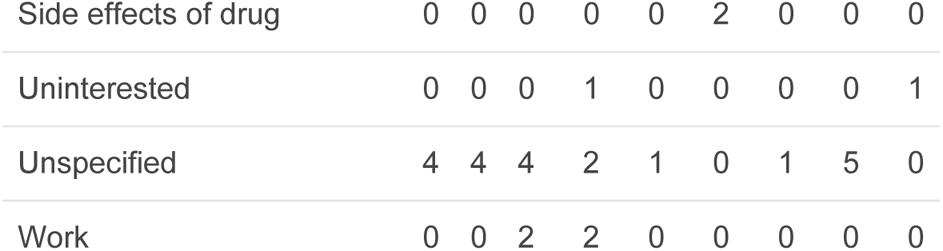

4 Withdrawals 4.1 Overall
|  | Days since enrollment |  |  |  |  |  |  |  |  |
| --- | --- | --- | --- | --- | --- | --- | --- | --- | --- |
|  | 3 | 4 | 5 | 10 | 11 | 14 | 15 | 21 | 28 |
| Family withdrew | 0 | 0 | 7 | 0 | 0 | 2 | 0 | 0 | 0 |
| Interpersonal problems | 0 | 4 | 3 | 0 | 0 | 0 | 0 | 0 | 0 |
| Lack of time | 0 | 1 | 0 | 0 | 0 | 0 | 0 | 0 | 0 |
| Other | 0 | 0 | 1 | 0 | 0 | 0 | 0 | 0 | 0 |
| Parental decision | 0 | 0 | 2 | 0 | 0 | 0 | 0 | 0 | 0 |
| Poor health | 0 | 0 | 1 | 0 | 0 | 1 | 0 | 0 | 0 |
| Sample collection problems | 0 | 0 | 1 | 0 | 0 | 0 | 0 | 0 | 0 |
| Side effects of drug | 0 | 0 | 0 | 0 | 0 | 2 | 0 | 0 | 0 |
| Uninterested | 0 | 0 | 0 | 1 | 0 | 0 | 0 | 0 | 1 |
| Unspecified | 4 | 4 | 4 | 2 | 1 | 0 | 1 | 5 | 0 |
| Work | 0 | 0 | 2 | 2 | 0 | 0 | 0 | 0 | 0 |

### 4.2 By Treatment Arm

| IFN |  |  |  |  |  |  |  |  |  | SOC |  |  |  |  |  |  |  |  |  |
| --- | --- | --- | --- | --- | --- | --- | --- | --- | --- | --- | --- | --- | --- | --- | --- | --- | --- | --- | --- |
|  | Days since enrollment |  |  |  |  |  |  |  |  |  | Days since enrollment |  |  |  |  |  |  |  |  |
|  | 3 | 4 | 5 | 10 | 11 | 14 | 15 | 21 | 28 |  | 3 | 4 | 5 | 10 | 11 | 14 | 15 | 21 | 28 |
| Family withdrew | 0 | 0 | 5 | 0 | 0 | 2 | 0 | 0 | 0 | Family withdrew | 0 | 0 | 2 | 0 | 0 | 0 | 0 | 0 | 0 |
| Interpersonal problems | 0 | 0 | 3 | 0 | 0 | 0 | 0 | 0 | 0 | Interpersonal problems | 0 | 4 | 0 | 0 | 0 | 0 | 0 | 0 | 0 |
| Lack of time | 0 | 1 | 0 | 0 | 0 | 0 | 0 | 0 | 0 | Lack of time | 0 | 0 | 0 | 0 | 0 | 0 | 0 | 0 | 0 |
| Other | 0 | 0 | 0 | 0 | 0 | 0 | 0 | 0 | 0 | Other | 0 | 0 | 1 | 0 | 0 | 0 | 0 | 0 | 0 |
| Parental decision | 0 | 0 | 2 | 0 | 0 | 0 | 0 | 0 | 0 | Parental decision | 0 | 0 | 0 | 0 | 0 | 0 | 0 | 0 | 0 |
| Poor health | 0 | 0 | 1 | 0 | 0 | 1 | 0 | 0 | 0 | Poor health | 0 | 0 | 0 | 0 | 0 | 0 | 0 | 0 | 0 |
| Sample collection problems | 0 | 0 | 0 | 0 | 0 | 0 | 0 | 0 | 0 | Sample collection problems | 0 | 0 | 1 | 0 | 0 | 0 | 0 | 0 | 0 |
| Side effects of drug | 0 | 0 | 0 | 0 | 0 | 2 | 0 | 0 | 0 | Side effects of drug | 0 | 0 | 0 | 0 | 0 | 0 | 0 | 0 | 0 |
| Uninterested | 0 | 0 | 0 | 1 | 0 | 0 | 0 | 0 | 1 | Uninterested | 0 | 0 | 0 | 0 | 0 | 0 | 0 | 0 | 0 |
| Unspecified | 4 | 3 | 1 | 2 | 1 | 0 | 0 | 0 | 0 | Unspecified | 0 | 1 | 3 | 0 | 0 | 0 | 1 | 5 | 0 |
| Work | 0 | 0 | 2 | 2 | 0 | 0 | 0 | 0 | 0 | Work | 0 | 0 | 0 | 0 | 0 | 0 | 0 | 0 | 0 |

All participants consented to the use of their collected data.

## 5 Missing Data

No imputation was performed.

## 6 Baseline Characteristics

### 6.1 Full Analysis Population

|  | Overall<br>(N=1172) |
| --- | --- |
| **Age (years)** |  |
| Mean (SD) | 34.1 (19.0) |
| **Sex** |  |
| Female | 617 (52.6%) |
| Male | 555 (47.4%) |
| **Household occupants** |  |
| Mean (SD) | 4.31 (1.73) |
| **Fully vaccinated for SARS-CoV-2** |  |
| No | 818 (69.8%) |
| Yes | 354 (30.2%) |
| **Cancer** |  |
| No | 1157 (98.7%) |
| Yes | 14 (1.2%) |
| Missing | 1 (0.1%) |
| **Diabetes** |  |
| No | 1125 (96.0%) |
| Yes | 46 (3.9%) |
| Missing | 1 (0.1%) |
| **Heart disease** |  |
| No | 1111 (94.8%) |
| Yes | 60 (5.1%) |
| Missing | 1 (0.1%) |
| **Asthma** |  |
| No | 1105 (94.3%) |
| Yes | 66 (5.6%) |
| Missing | 1 (0.1%) |
| **Chronic lung disease** |  |
| No | 1170 (99.8%) |
| Yes | 1 (0.1%) |
| Missing | 1 (0.1%) |
| **Chronic kidney disease** |  |
| No | 1168 (99.7%) |
| Yes | 3 (0.3%) |
| Missing | 1 (0.1%) |
| **Chronic neurological disease** |  |
| No | 1164 (99.3%) |
| Yes | 7 (0.6%) |
| Missing | 1 (0.1%) |
| **Smoker** |  |
| No | 1021 (87.1%) |
| Yes | 151 (12.9%) |

6.2 IC-ITT Population
|  | SOC<br>(N=169) | IFN<br>(N=172) |
| --- | --- | --- |
| **Age (years)** |  |  |
| Mean (SD) | 41.3 (12.2) | 40.4 (12.8) |
| **Sex** |  |  |
| Female | 82 (48.5%) | 79 (45.9%) |
| Male | 87 (51.5%) | 93 (54.1%) |
| **Household occupants** |  |  |
| Mean (SD) | 3.74 (1.29) | 3.90 (1.64) |
| **Fully vaccinated for SARS-CoV-2** |  |  |
| No | 106 (62.7%) | 116 (67.4%) |
| Yes | 63 (37.3%) | 56 (32.6%) |
| **Cancer** |  |  |
| No | 166 (98.2%) | 169 (98.3%) |
| Yes | 3 (1.8%) | 3 (1.7%) |
| **Diabetes** |  |  |
| No | 162 (95.9%) | 163 (94.8%) |
| Yes | 7 (4.1%) | 9 (5.2%) |
| **Heart disease** |  |  |
| No | 158 (93.5%) | 165 (95.9%) |
| Yes | 11 (6.5%) | 7 (4.1%) |
| **Asthma** |  |  |
| No | 157 (92.9%) | 167 (97.1%) |
| Yes | 12 (7.1%) | 5 (2.9%) |
| **Chronic lung disease** |  |  |
| No | 169 (100%) | 172 (100%) |
| Yes | 0 (0%) | 0 (0%) |
| **Chronic kidney disease** |  |  |
| No | 169 (100%) | 172 (100%) |
| Yes | 0 (0%) | 0 (0%) |
| **Chronic neurological disease** |  |  |
| No | 169 (100%) | 172 (100%) |
| Yes | 0 (0%) | 0 (0%) |
| **Smoker** |  |  |
| No | 147 (87.0%) | 138 (80.2%) |
| Yes | 22 (13.0%) | 34 (19.8%) |

### 6.2 IC-ITT Population

6.3 IC-INF Population
|  | **SOC**<br>**(N=250)** | **IFN**<br>**(N=252)** |
| --- | --- | --- |
| **Age (years)** |  |  |
| Mean (SD) | 42.2 (13.1) | 40.5 (13.5) |
| **Sex** |  |  |
| Female | 129 (51.6%) | 117 (46.4%) |
| Male | 121 (48.4%) | 135 (53.6%) |
| **Household occupants** |  |  |
| Mean (SD) | 3.83 (1.28) | 3.98 (1.65) |
| **Fully vaccinated for SARS-CoV-2** |  |  |
| No | 158 (63.2%) | 170 (67.5%) |
| Yes | 92 (36.8%) | 82 (32.5%) |
| **Cancer** |  |  |
| No | 246 (98.4%) | 248 (98.4%) |
| Yes | 4 (1.6%) | 4 (1.6%) |
| **Diabetes** |  |  |
| No | 237 (94.8%) | 237 (94.0%) |
| Yes | 13 (5.2%) | 15 (6.0%) |
| **Heart disease** |  |  |
| No | 232 (92.8%) | 236 (93.7%) |
| Yes | 18 (7.2%) | 16 (6.3%) |
| **Asthma** |  |  |
| No | 230 (92.0%) | 239 (94.8%) |
| Yes | 20 (8.0%) | 13 (5.2%) |
| **Chronic lung disease** |  |  |
| No | 250 (100%) | 252 (100%) |
| Yes | 0 (0%) | 0 (0%) |
| **Chronic kidney disease** |  |  |
| No | 250 (100%) | 252 (100%) |
| Yes | 0 (0%) | 0 (0%) |
| **Chronic neurological disease** |  |  |
| No | 249 (99.6%) | 251 (99.6%) |
| Yes | 1 (0.4%) | 1 (0.4%) |
| **Smoker** |  |  |
| No | 215 (86.0%) | 205 (81.3%) |
| Yes | 35 (14.0%) | 47 (18.7%) |

### 6.3 IC-INF Population

6.4 EHC-ITT Population
|  | **SOC**<br>**(N=165)** | **IFN**<br>**(N=177)** |
| --- | --- | --- |
| **Age (years)** |  |  |
| Mean (SD) | 41.6 (15.5) | 38.0 (14.8) |
| **Sex** |  |  |
| Female | 93 (56.4%) | 107 (60.5%) |
| Male | 72 (43.6%) | 70 (39.5%) |
| **Household occupants** |  |  |
| Mean (SD) | 4.00 (1.37) | 4.54 (2.18) |
| **Fully vaccinated for SARS-CoV-2** |  |  |
| No | 80 (48.5%) | 107 (60.5%) |
| Yes | 85 (51.5%) | 70 (39.5%) |
| **Cancer** |  |  |
| No | 165 (100%) | 176 (99.4%) |
| Yes | 0 (0%) | 1 (0.6%) |
| **Diabetes** |  |  |
| No | 160 (97.0%) | 169 (95.5%) |
| Yes | 5 (3.0%) | 8 (4.5%) |
| **Heart disease** |  |  |
| No | 160 (97.0%) | 170 (96.0%) |
| Yes | 5 (3.0%) | 7 (4.0%) |
| **Asthma** |  |  |
| No | 157 (95.2%) | 170 (96.0%) |
| Yes | 8 (4.8%) | 7 (4.0%) |
| **Chronic lung disease** |  |  |
| No | 165 (100%) | 177 (100%) |
| Yes | 0 (0%) | 0 (0%) |
| **Chronic kidney disease** |  |  |
| No | 165 (100%) | 177 (100%) |
| Yes | 0 (0%) | 0 (0%) |
| **Chronic neurological disease** |  |  |
| No | 165 (100%) | 175 (98.9%) |
| Yes | 0 (0%) | 2 (1.1%) |
| **Smoker** |  |  |
| No | 140 (84.8%) | 145 (81.9%) |
| Yes | 25 (15.2%) | 32 (18.1%) |

### 6.4 EHC-ITT Population

6.5 HC Population
|  | **SOC**<br>**(N=315)** | **IFN**<br>**(N=355)** |
| --- | --- | --- |
| **Age (years)** |  |  |
| Mean (SD) | 29.3 (21.5) | 28.0 (20.2) |
| **Sex** |  |  |
| Female | 165 (52.4%) | 206 (58.0%) |
| Male | 150 (47.6%) | 149 (42.0%) |
| **Household occupants** |  |  |
| Mean (SD) | 4.31 (1.29) | 4.89 (2.18) |
| **Fully vaccinated for SARS-CoV-2** |  |  |
| No | 222 (70.5%) | 268 (75.5%) |
| Yes | 93 (29.5%) | 87 (24.5%) |
| **Cancer** |  |  |
| No | 313 (99.4%) | 350 (98.6%) |
| Yes | 1 (0.3%) | 5 (1.4%) |
| Missing | 1 (0.3%) | 0 (0%) |
| **Diabetes** |  |  |
| No | 305 (96.8%) | 346 (97.5%) |
| Yes | 9 (2.9%) | 9 (2.5%) |
| Missing | 1 (0.3%) | 0 (0%) |
| **Heart disease** |  |  |
| No | 301 (95.6%) | 342 (96.3%) |
| Yes | 13 (4.1%) | 13 (3.7%) |
| Missing | 1 (0.3%) | 0 (0%) |
| **Asthma** |  |  |
| No | 294 (93.3%) | 342 (96.3%) |
| Yes | 20 (6.3%) | 13 (3.7%) |
| Missing | 1 (0.3%) | 0 (0%) |
| **Chronic lung disease** |  |  |
| No | 314 (99.7%) | 354 (99.7%) |
| Yes | 0 (0%) | 1 (0.3%) |
| Missing | 1 (0.3%) | 0 (0%) |
| **Chronic kidney disease** |  |  |
| No | 314 (99.7%) | 352 (99.2%) |
| Yes | 0 (0%) | 3 (0.8%) |
| Missing | 1 (0.3%) | 0 (0%) |
| **Chronic neurological disease** |  |  |
| No | 313 (99.4%) | 351 (98.9%) |
| Yes | 1 (0.3%) | 4 (1.1%) |
| Missing | 1 (0.3%) | 0 (0%) |
| **Smoker** |  |  |
| No | 287 (91.1%) | 314 (88.5%) |
| Yes | 28 (8.9%) | 41 (11.5%) |

### 6.5 HC Population

| Covariate | OR | 95% CI |
| --- | --- | --- |
| Intercept | 0.199 | [0.0284, 0.8756] |
| Age (bin 2) | 1.519 | [ 0.342, 10.616] |
| Age (bin 3) | 1.129 | [0.254, 7.891] |
| Age (bin 4) | 1.504 | [ 0.336, 10.552] |
| Male | 2.138 | [1.414, 3.263] |
| IFN treatment | 0.979 | [0.647, 1.479] |

## 7 Primary Analyses SAP 6.10

### 7.1 Primary Analysis 1 SAP 6.10.1

#### 7.1.1 Main Analysis

This corresponds to SAP section 6.10.1 (Primary Analysis 1). Primary analysis 1 is carried out on participants from the IC-INF population, to compare the probability of index cases being SARS-CoV-2 negative vs positive per saliva PCR on the study day 11 saliva sample (“Not Shedding vs. Shedding”) in either the treatment or soc arm of the study.

A total of 468 participants were used in this analysis.

| Treatment | N | AR | ARR | 95% CI |
| --- | --- | --- | --- | --- |
| SOC | 237 | 0.287 |  |  |
| IFN | 225 | 0.289 | -0.00197 | [-0.0846, 0.0806] |

#### 7.1.2 Supporting Analyses

##### 7.1.2.1 Absolute Risk per Treatment Arm

A total of 468 participants were used in this analysis.

| Covariate | OR | 95% CI |
| --- | --- | --- |
| Intercept | 0.202 | [0.0289, 0.8893] |
| Age (bin 2) | 1.654 | [ 0.372, 11.570] |
| Age (bin 3) | 1.352 | [0.302, 9.494] |
| Age (bin 4) | 1.884 | [ 0.416, 13.333] |
| Male | 2.119 | [1.398, 3.241] |
| Vaccinated | 0.604 | [0.376, 0.956] |
| IFN treatment | 0.963 | [0.635, 1.459] |

##### 7.1.2.2 GLM with a Log Link

A total of 468 participants were used in this analysis.

This analysis did not converge.

#### 7.1.3 Sensitivity Analyses

##### 7.1.3.1 Adjusted for Vaccination Status

A total of 468 participants were used in this analysis.

| Covariate | OR | 95% CI |
| --- | --- | --- |
| Intercept | 0.115 | [0.00566, 0.81281] |
| Age (bin 2) | 1.961 | [ 0.284, 39.261] |
| Age (bin 3) | 1.272 | [ 0.184, 25.425] |
| Age (bin 4) | 2.453 | [ 0.351, 49.321] |
| Male | 2.224 | [1.315, 3.833] |
| IFN treatment | 1.106 | [0.657, 1.863] |

##### 7.1.3.2 IC-ITT Population

This corresponds to SAP section 6.10.1 (sensitivity analysis B). Sentivity analysis B is carried out on participants from the IC-ITT population.

A total of 323 participants were used in this analysis.

| Covariate | OR | 95% CI |
| --- | --- | --- |
| Intercept | 0.122 | [0.006, 0.865] |
| Age (bin 2) | 1.935 | [ 0.280, 38.738] |
| Age (bin 3) | 1.178 | [ 0.169, 23.638] |
| Age (bin 4) | 2.361 | [ 0.337, 47.531] |
| Male | 2.156 | [1.259, 3.760] |
| IFN treatment | 1.062 | [0.622, 1.810] |

##### 7.1.3.3 IC-PP Population

This corresponds to SAP section 6.10.1 (sensitivity analysis C). Sentivity analysis C is carried out on participants from the IC-PP population (excluding participants randomized to the IFN arm that received <3 doses; shown below).

A total of 310 participants were used in this analysis.

| Covariate | OR | 95% CI |
| --- | --- | --- |
| Intercept | 0.294 | [0.0246, 2.7823] |
| Age (bin 2) | 1.519 | [ 0.342, 10.629] |
| Age (bin 3) | 1.138 | [0.256, 7.959] |
| Age (bin 4) | 1.539 | [ 0.343, 10.807] |
| Male | 2.114 | [1.397, 3.229] |
| Days between symptom onset and visit date | 0.973 | [0.866, 1.090] |
| IFN treatment | 0.987 | [0.652, 1.494] |

#### 7.1.4 Unplanned Analyses

##### 7.1.4.1 Adjusted for Days between Symptom Onset and Visit

A total of 468 participants were used in this analysis.

This is an unplanned analysis. Days between symptom onset and visit date was included as a covariate.

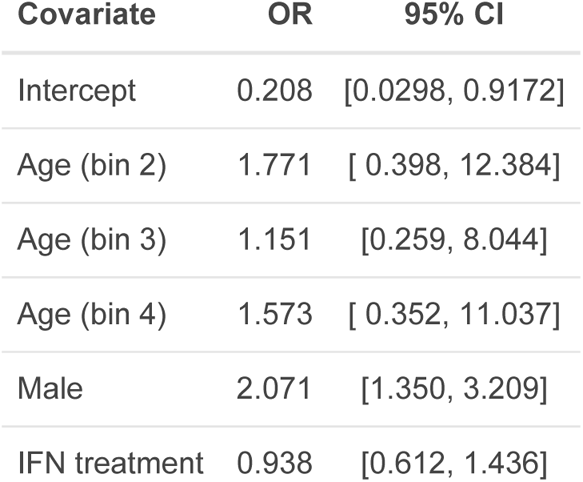

##### 7.1.4.2 Excluding PCR Negative Index cases

A total of 427 participants were used in this analysis.

This is an unplanned analysis. Index cases that were PCR negative at both study day 1 and study day 6 were excluded.

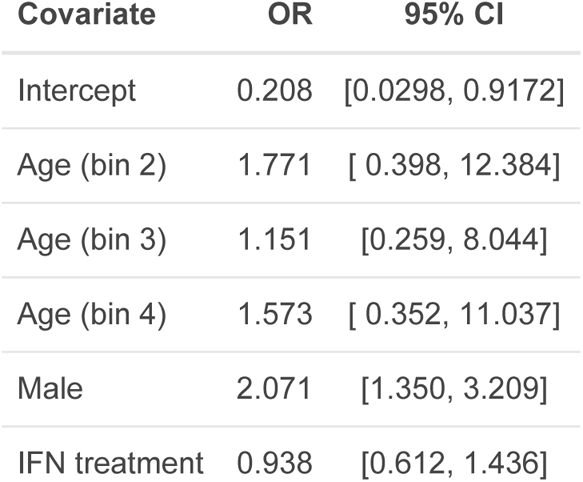

Below, I repeat the above, excluding ***households*** where the index case was PCR negative at both study day 1 and 6.

| Covariate | OR | 95% CI |
| --- | --- | --- |
| Intercept | 0.229 | [0.0319, 1.0685] |
| Age (bin 2) | 1.708 | [ 0.364, 12.208] |
| Age (bin 3) | 1.227 | [0.260, 8.789] |
| Age (bin 4) | 1.798 | [ 0.376, 12.999] |
| Male | 2.128 | [1.383, 3.307] |
| Vaccinated | 0.573 | [0.349, 0.924] |
| IFN treatment | 0.953 | [0.619, 1.466] |

### 7.2 Primary Analysis 2 SAP 6.10.2

#### 7.2.1 Main Analysis

This corresponds to SAP section 6.10.2 (primary analysis 2). Primary analysis 2 is carried out on participants from the EHC-ITT population, to compare the probability of eligible household contacts being SARS-CoV-2 negative vs positive per saliva PCR on study day 11 (“Not Shedding vs. Shedding”) in either the treatment or soc arm of the study.

A total of 322 participants were used in this analysis.

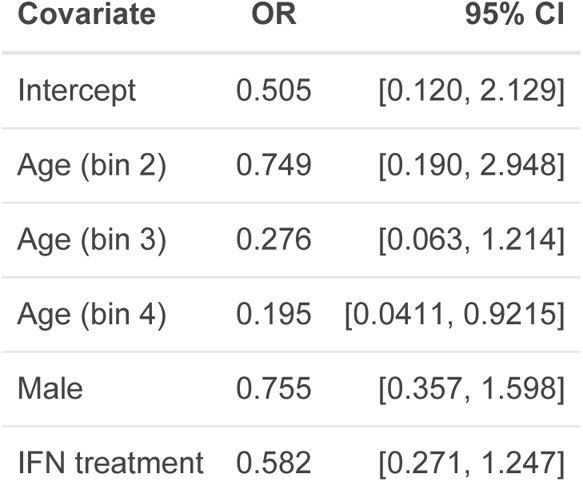

| Covariate | OR | 95% CI |
| --- | --- | --- |
| Intercept | 0.505 | [0.120, 2.129] |
| Age (bin 2) | 0.749 | [0.190, 2.948] |
| Age (bin 3) | 0.276 | [0.063, 1.214] |
| Age (bin 4) | 0.195 | [0.0411, 0.9215] |
| Male | 0.755 | [0.357, 1.598] |
| IFN treatment | 0.582 | [0.271, 1.247] |

#### 7.2.2 Supporting Analyses

##### 7.2.2.1 Absolute Risk per Treatment Arm

A total of 322 participants were used in this analysis.

| Treatment | N | AR | ARR | 95% CI |
| --- | --- | --- | --- | --- |
| SOC | 156 | 0.154 |  |  |
| IFN | 165 | 0.115 | 0.0387 | [-0.036, 0.113] |

##### 7.2.2.2 GLMM with a Log Link

A total of 322 participants were used in this analysis.

This analysis did not converge.

#### 7.2.3 Sensitivity Analyses

##### 7.2.3.1 Adjusted for Risk Factors

A total of 322 participants were used in this analysis.

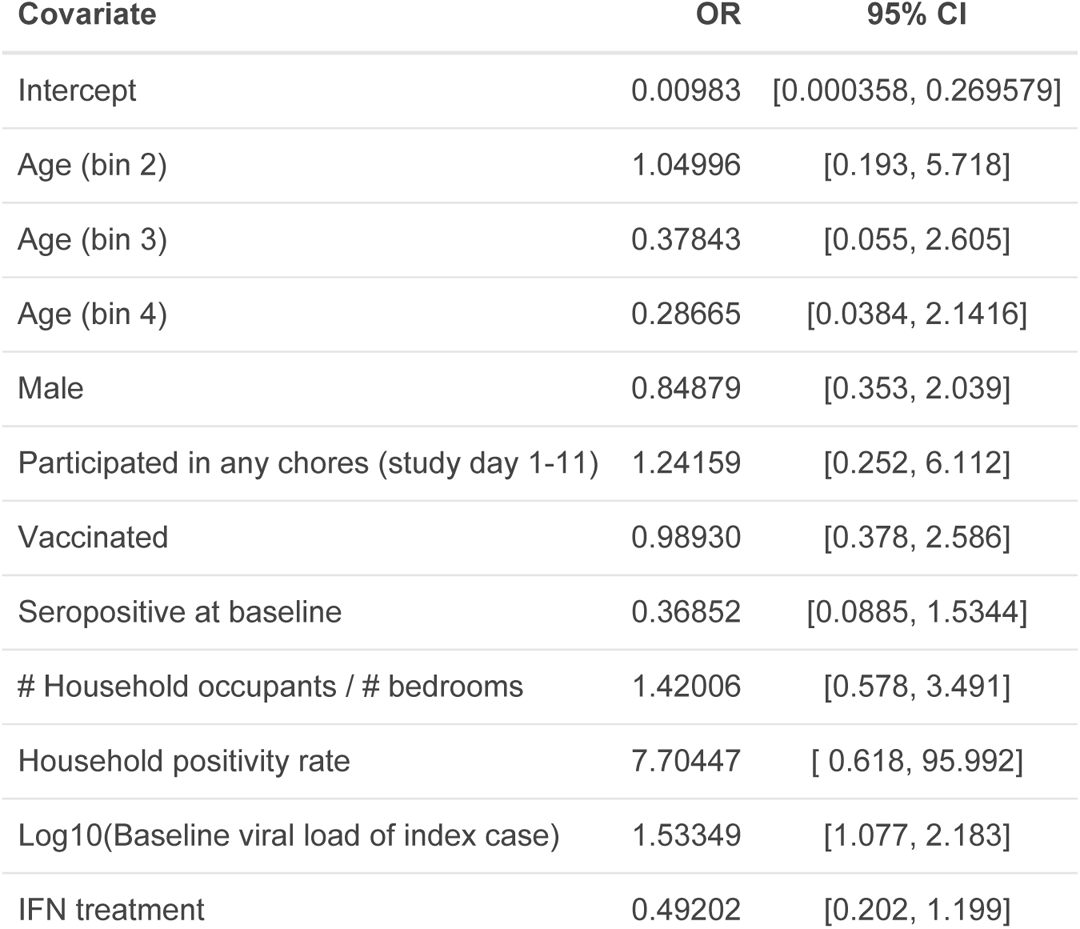

##### 7.2.3.2 HC Population

A total of 633 participants were used in this analysis.

7.2.3.2 HC Population A total of 633 participants were used in this analysis.
| Covariate | OR | 95% CI |
| --- | --- | --- |
| Intercept | 0.194 | [0.0679, 0.5543] |
| armNE | 2.274 | [0.917, 5.640] |
| Age (bin 2) | 1.432 | [0.499, 4.109] |
| Age (bin 3) | 0.883 | [0.292, 2.665] |
| Age (bin 4) | 0.314 | [0.105, 0.941] |
| Male | 0.818 | [0.464, 1.443] |
| IFN treatment | 0.577 | [0.287, 1.160] |

##### 7.2.3.3 EHC-PP Population

A total of 312 participants were used in this analysis.

7.2.3.3 EHC-PP Population A total of 312 participants were used in this analysis.
| Covariate | OR | 95% CI |
| --- | --- | --- |
| Intercept | 0.574 | [0.133, 2.487] |
| Age (bin 2) | 0.670 | [0.166, 2.705] |
| Age (bin 3) | 0.257 | [0.0572, 1.1555] |
| Age (bin 4) | 0.174 | [0.0359, 0.8411] |
| Male | 0.695 | [0.323, 1.495] |
| IFN treatment | 0.589 | [0.272, 1.276] |

#### 7.2.4 Unplanned Analyses

##### 7.2.4.1 Adjusted for Days between Symptom Onset and Visit

A total of 322 participants were used in this analysis.

This is an unplanned analysis. Days between symptom onset *of the index case* and visit date was included as a covariate. The analysis did not converge.

## 8 Secondary Analyses

### 8.1 Secondary Analysis 1 SAP 6.10.3

This corresponds to SAP section 6.10.3 (secondary analysis 1). This analysis is carried out on the IC-INF population, using discrete time survival analysis to assess whether IFN treatment has an effect on the duration for which participants had SARS-CoV-2 positive saliva PCR.

Note that a higher hazard ratio is better, as it indicates a faster time to discharge from hospital.

#### 8.1.1 Main Analysis

A total of 502 participants (totalling 2807 visits) were used in this analysis.

| Covariate | HR | 95% CI |
| --- | --- | --- |
| Intercept | 0.286 | [0.179, 0.443] |
| Age (bin 2) | 0.836 | [0.540, 1.330] |
| Age (bin 3) | 0.799 | [0.517, 1.271] |
| Age (bin 4) | 0.833 | [0.537, 1.327] |
| Male | 0.818 | [0.728, 0.918] |
| Days since enrollment | 1.110 | [1.102, 1.118] |
| IFN treatment | 1.083 | [0.964, 1.217] |

#### 8.1.2 Sensitivity Analyses

##### 8.1.2.1 IC-ITT Population

A total of 341 participants (totalling 1927 visits) were used in this analysis.

| Covariate | HR | 95% CI |
| --- | --- | --- |
| Intercept | 0.338 | [0.190, 0.576] |
| Age (bin 2) | 0.830 | [0.491, 1.460] |
| Age (bin 3) | 0.800 | [0.474, 1.405] |
| Age (bin 4) | 0.686 | [0.403, 1.213] |
| Male | 0.888 | [0.773, 1.021] |
| Days since enrollment | 1.108 | [1.099, 1.118] |
| IFN treatment | 1.018 | [0.886, 1.171] |

##### 8.1.2.2 IC-PP Population

A total of 315 participants (totalling 1830 visits) were used in this analysis.

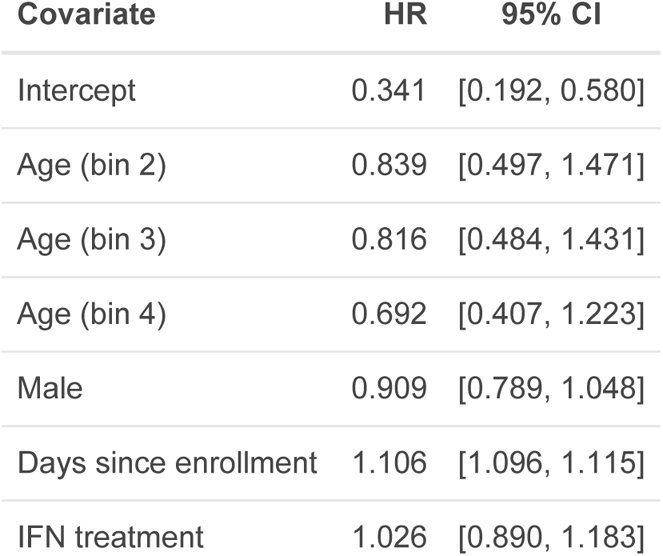

8.1.2.2 IC-PP Population A total of 315 participants (totalling 1830 visits) were used in this analysis.
| Covariate | HR | 95% CI |
| --- | --- | --- |
| Intercept | 0.341 | [0.192, 0.580] |
| Age (bin 2) | 0.839 | [0.497, 1.471] |
| Age (bin 3) | 0.816 | [0.484, 1.431] |
| Age (bin 4) | 0.692 | [0.407, 1.223] |
| Male | 0.909 | [0.789, 1.048] |
| Days since enrollment | 1.106 | [1.096, 1.115] |
| IFN treatment | 1.026 | [0.890, 1.183] |

### 8.2 Secondary Analysis 2 SAP 6.10.4

This corresponds to SAP section 6.10.4 (secondary analysis 2). This analysis is carried out on participants from the HC population.

#### 8.2.1 Main Analysis

##### 8.2.1.1 Incidence of SARS-CoV-2 Positive Saliva PCR on Study Day 1

The HC population consists of only household contacts that were PCR negative at visit 1, so this comparison is not applicable.

##### 8.2.1.2 Incidence of SARS-CoV-2 Positive Saliva PCR on Study Day 11

A total of 633 participants were used in this analysis.

| Incidence (SOC) | Incidence (IFN) | Cochran-Mantel-Haenszel p-value |
| --- | --- | --- |
| 0.208 | 0.173 | 0.329 |

##### 8.2.1.3 Seroconversion at Study Day 29

A total of 594 participants were used in this analysis.

This analysis is comparing the proportion of seropositive participants at study day 29, stratified by baseline immune status.

| Incidence (SOC) | Incidence (IFN) | Cochran-Mantel-Haenszel p-value |
| --- | --- | --- |
| 0.537 | 0.415 | 0.0558 |

### 8.3 Secondary Analysis 3 SAP 6.10.5

This corresponds to SAP section 6.10.5 (secondary analysis 3). This analysis is carried out on participants from the IC-ITT population.

For this analysis, households with size greater than 8 were combined with the households of size 8, as there were not many of the former.

#### 8.3.1 Main Analysis

##### 8.3.1.1 Incidence of Hospitalization Due to COVID-19

A total of 299 participants were used in this analysis.

| Outcome | Overall incidence in SOC | Overall incidence in IFN | Cochran-Mantel-Haenszel p-value |
| --- | --- | --- | --- |
| Hospitalized | 0.101 | 0.0938 | 0.820 |

##### 8.3.1.2 Incidence of Death Due to COVID-19

No deaths occurred in this analysis population.

##### 8.3.1.3 Incidence of Hospitalization and/or Death Due to COVID-19

As there were no deaths in this analysis population, the results are the same as “Incidence of Hospitalization Due to COVID-19”.

##### 8.3.1.4 Duration of Hospital Stay Due to COVID-19

A total of 29 participants were used in this analysis.

As there were no deaths in this analysis population, a Cox proportional-hazards model was used instead. Note that a higher hazard ratio is better, in this case, as it indicates a faster discharge from the hospital.

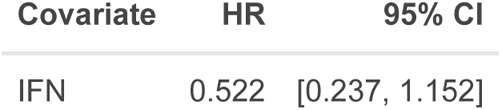

#### 8.3.2 Sensitivity Analyses

##### 8.3.2.1 IC-INF Population

**8.3.2.1.1 Incidence of Hospitalization Due to COVID-19**

A total of 437 participants were used in this analysis.

| Outcome | Overall incidence in SOC | Overall incidence in IFN | Cochran-Mantel-Haenszel p-value |
| --- | --- | --- | --- |
| Hospitalized | 0.109 | 0.0929 | 0.570 |

**8.3.2.1.2 Incidence of Death Due to COVID-19**

No deaths occurred in this analysis population.

**8.3.2.1.3 Incidence of Hospitalization and/or Death Due to COVID-19**

As there were no deaths in this analysis population, the results are the same as “Incidence of Hospitalization Due to COVID-19”.

**8.3.2.1.4 Duration of Hospital Stay Due to COVID-19**

A total of 43 participants were used in this analysis.

As there were no deaths in this analysis population, a Cox proportional-hazards model was used instead. Note that a higher hazard ratio is better, in this case, as it indicates a faster discharge from the hospital.

| Covariate | HR | 95% CI |
| --- | --- | --- |
| IFN | 0.452 | [0.233, 0.877] |

**Note discrepancy in number of hospitalizations reported (n = 44 vs. 43 reported) is caused by 1 participant where hospital duration could not be derived (CE 183 04 - no discharge date)**

##### 8.3.2.2 IC-PP Population

**8.3.2.2.1 Incidence of Hospitalization Due to COVID-19**

A total of 275 participants were used in this analysis.

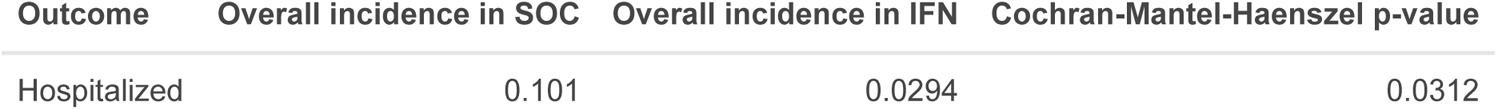

**8.3.2.2.2 Incidence of Death Due to COVID-19**

No deaths occurred in this analysis population.

**8.3.2.2.3 Incidence of Hospitalization and/or Death Due to COVID-19**

As there were no deaths in this analysis population, the results are the same as “Incidence of Hospitalization Due to COVID-19”.

**8.3.2.2.4 Duration of Hospital Stay Due to COVID-19**

A total of 18 participants were used in this analysis.

As there were no deaths in this analysis population, a Cox proportional-hazards model was used instead. Note that a higher hazard ratio is better, in this case, as it indicates a faster discharge from the hospital.

| Covariate | HR | 95% CI |
| --- | --- | --- |
| IFN | 2.006 | [0.601, 6.690] |

### 8.4 Secondary Analysis 4 SAP 6.10.6

#### 8.4.1 Main Analysis

A total of 844 participants (totalling 6784 visits) were used in this analysis.

##### 8.4.1.1 Incidence of Adverse Events

| Treatment | Rate of AE per participant | Incidence of any AE | Fisher's exact test |
| --- | --- | --- | --- |
| SOC | 1.824 | 0.280 |  |
| IFN | 2.117 | 0.235 | 0.156 |

##### 8.4.1.2 Incidence of Adverse Events Related To Treatment

| Treatment | Rate of related AE per participant | Incidence of any related AE | Fisher's exact test |
| --- | --- | --- | --- |
| SOC | 0.00964 | 0.00964 |  |
| IFN | 0.287 | 0.235 | 2.31e-27 |

##### 8.4.1.3 Incidence of Serious Adverse Events

| Treatment | Rate of SAE per participant | Incidence of any SAE | Fisher's exact test |
| --- | --- | --- | --- |
| SOC | 0.0627 | 0.0578 |  |
| IFN | 0.0559 | 0.0559 | 1.000 |

## 9 Safety Analysis SAP 6.11

See Secondary Analysis 4.

## 10 Additional Analyses SAP 6.12

### 10.1 Dose-response and Recovery SAP 6.12.1

|  | PCR negative at study day 11 | PCR positive at study day 11 |
| --- | --- | --- |
| 1 dose | 3 | 1 |
| 2 doses | 5 | 3 |
| 3 doses | 109 | 37 |

The p-value for the Fisher’s exact test is 0.756.

### 10.2 Dose-response and Transmission SAP 6.12.2

|  | PCR negative at study day 11 | PCR positive at study day 11 |
| --- | --- | --- |
| 1 dose | 6 | 0 |
| 2 doses | 3 | 1 |
| 3 doses | 137 | 18 |

The p-value for the Fisher’s exact test is 0.481.

### 10.3 Viral Load SAP 6.12.3

Growth rate is undefined (i.e. is missing) for participants whose peak viral load occurred on study day 1. Duration of infection is undefined. for participants whose last PCR result was positive. Decay is undefined for participants if their duration of infection is undefined.

|  | SOC<br>(N=209) | IFN<br>(N=209) |
| --- | --- | --- |
| **Peak viral load** |  |  |
| Mean (SD) | 28,913,933 (196,793,258) | 6,591,476 (33,775,900) |
| 95% CI | [-364,672,583; 422,500,449] | [-60,960,324; 74,143,277] |
| Median [Q1; Q3] | 220,461 [28,079; 1,741,659] | 220,461 [32,374; 1,411,196] |
| **Days to peak viral load** |  |  |
| Mean (SD) | 1.87 (4.52) | 1.40 (3.02) |
| 95% CI | [-7.17; 10.92] | [-4.64; 7.44] |
| Median [Q1; Q3] | 0.00 [0.00; 0.00] | 0.00 [0.00; 0.00] |
| **Growth rate** |  |  |
| Mean (SD) | 262,111 (951,800) | 431,891 (1,431,385) |
| 95% CI | [-1,641,490; 2,165,711] | [-2,430,878; 3,294,661] |
| Median [Q1; Q3] | 15,083 [2,003; 100,240] | 4,747 [456; 40,673] |
| Missing | 163 (78.0%) | 163 (78.0%) |
| **Duration of infection** |  |  |
| Mean (SD) | 8.88 (6.89) | 8.38 (6.29) |
| 95% CI | [-4.90; 22.66] | [-4.19; 20.95] |
| Median [Q1; Q3] | 6.00 [1.00; 16.00] | 6.00 [6.00; 11.00] |
| Missing | 31 (14.8%) | 26 (12.4%) |
| **Decay rate** |  |  |
| Mean (SD) | 1,376,272 (4,574,811) | 948,980 (4,379,841) |
| 95% CI | [-7,773,350; 10,525,894] | [-7,810,702; 9,708,662] |
| Median [Q1; Q3] | 58,102 [10,146; 294,797] | 43,171 [11,186; 268,371] |
| Missing | 33 (15.8%) | 26 (12.4%) |
| **Area under the curve** |  |  |
| Mean (SD) | 75,272,876 (494,951,264) | 19,688,768 (91,915,780) |
| 95% CI | [-914,629,653; 1,065,175,405] | [-164,142,792; 203,520,328] |
| Median [Q1; Q3] | 698,934 [103,353; 6,628,988] | 782,976 [127,664; 3,900,093] |
| Missing | 2 (1.0%) | 0 (0%) |
| **Baseline viral load** |  |  |
| Mean (SD) | 28,590,599 (196,824,913) | 6,090,935 (33,614,342) |
| 95% CI | [-365,059,226; 422,240,425] | [-61,137,749; 73,319,620] |
| Median [Q1; Q3] | 144,737 [10,562; 1,270,280] | 133,549 [18,435; 870,885] |

Repeat including p-values for the comparisons between the groups (Wilcoxon Rank Sum Test).

|  | SOC<br>(N=209) | IFN<br>(N=209) | P |
| --- | --- | --- | --- |
| Peak viral load |  |  |  |
| Mean (SD) | 28,913,933 (196,793,258) | 6,591,476 (33,775,900) | 0.761 |
| 95% CI | [-364,672,583; 422,500,449] | [-60,960,324; 74,143,277] |  |
| Median [Q1; Q3] | 220,461 [28,079; 1,741,659] | 220,461 [32,374; 1,411,196] |  |
| Days to peak viral load |  |  |  |
| Mean (SD) | 1.87 (4.52) | 1.40 (3.02) | 0.855 |
| 95% CI | [-7.17; 10.92] | [-4.64; 7.44] |  |
| Median [Q1; Q3] | 0.00 [0.00; 0.00] | 0.00 [0.00; 0.00] |  |
| Growth rate |  |  |  |
| Mean (SD) | 262,111 (951,800) | 431,891 (1,431,385) | 0.416 |
| 95% CI | [-1,641,490; 2,165,711] | [-2,430,878; 3,294,661] |  |
| Median [Q1; Q3] | 15,083 [2,003; 100,240] | 4,747 [456; 40,673] |  |
| Duration of infection |  |  |  |
| Mean (SD) | 8.88 (6.89) | 8.38 (6.29) | 0.618 |
| 95% CI | [-4.90; 22.66] | [-4.19; 20.95] |  |
| Median [Q1; Q3] | 6.00 [1.00; 16.00] | 6.00 [6.00; 11.00] |  |
| Decay rate |  |  |  |
| Mean (SD) | 1,376,272 (4,574,811) | 948,980 (4,379,841) | 0.59 |
| 95% CI | [-7,773,350; 10,525,894] | [-7,810,702; 9,708,662] |  |
| Median [Q1; Q3] | 58,102 [10,146; 294,797] | 43,171 [11,186; 268,371] |  |
| Area under the curve |  |  |  |
| Mean (SD) | 75,272,876 (494,951,264) | 19,688,768 (91,915,780) | 0.856 |
| 95% CI | [-914,629,653; 1,065,175,405] | [-164,142,792; 203,520,328] |  |
| Median [Q1; Q3] | 698,934 [103,353; 6,628,988] | 782,976 [127,664; 3,900,093] |  |
| Baseline viral load |  |  |  |
| Mean (SD) | 28,590,599 (196,824,913) | 6,090,935 (33,614,342) | 0.883 |
| 95% CI | [-365,059,226; 422,240,425] | [-61,137,749; 73,319,620] |  |
| Median [Q1; Q3] | 144,737 [10,562; 1,270,280] | 133,549 [18,435; 870,885] |  |

## 11 Optional Analyses

### 11.1 Bayesian Analysis

This analysis was not performed.

### 11.2 Exploratory Two-period Generalized Mixed Effects Model

This analysis is detailed in a separate document.

## Supplementary Appendices B

### Containing Coronavirus Disease 19 Trial (CONCORD-19): Exploratory Analyses

Exploratory analysis, two-period glmer to explore treatment effect during and after treatment period in study-eligible and ineligible household contacts.

Casey P. Shannon (PROOF Centre of Excellence, University of British Columbia), true, Robert Balshaw (Centre for Healthcare Innovation, University of Manitoba)

2022-05-19

### Introduction

Primary analysis 2 was carried out on participants from the EHC-ITT population, to compare the probability of eligible household contacts being SARS-CoV-2 negative vs positive per saliva PCR on study day 11 (“Not Shedding vs. Shedding”) in either the treatment or soc arm of the study.

A planned sensitivity analysis adjusting for various risk factors had promising results (reduction of risk in the IFN arm), though this effect did not reach the pre-specified statistical significance threshold.

**Here we revisit primary analysis 2, adopting a slightly modified approach incorporating treatment periods, and additionally considering non-eligible household contacts.**

### 1. New outcome measure

Rather than counting participants as positive based on their study day 11 sample SARS-CoV-2 saliva PCR, we count participants as PCR-positive according to whether they have had their first positive SARS-CoV-2 saliva PCR on any study day during period 1 (**during treatment and isolation; D1-D11 inclusive**) or period 2 (**after treatment and isolation; D12-D29**), or not at all.

Participants who test positive in period 1 are *not* counted in period two. Effectively, they are censored at their first positive saliva PCR as they are no longer at risk of becoming positive for the first time.

### 2. Inclusion of additional covariates

**Household-level (hh_cov)**

- Number of individuals in the household
- Number of vaccinated individuals in the household
- Viral load (log10 viral count) of the household index case on study day 1
- Number of individuals in the household with positive saliva PCR *by* the start of the period

**These household level counts consider all household members (index cases and all treatment-eligible and treatment ineligible individuals in the household).**

**Individual-level (indv_cov)**

- Participant eligibility (CE; eligible household contact/NE; non-eligible household contact)
- Participant age
- Participant sex

### 3. Models

- pcr ~ (1 | household) + hh_cov + indv_cov + period
- pcr ~ (1 | household) + hh_cov + indv_cov + period + trt
- pcr ~ (1 | household) + hh_cov + indv_cov + period + trt + period:trt

We test the hypothesis of an overall treatment effect by comparing model 2 to model 1 using the likelihood ratio test (LRT).

We test the hypothesis of an interaction between period and treatment (i.e., does treatment effect differ between the two periods) by comparing model 3 to model 2 using the likelihood ratio test (LRT).

**P-values less than 0.05 are described as “significant”, as is conventional, but the post-hoc nature of this analysis must be remembered.**

### 4. Defining eligibility

Following internal discussion, we elected to exclude households from the analysis if:

- The index case subsequently tested negative (by saliva PCR) on both study day 1 AND study day 6 (negative index case)
- There were no negative household members (eligible OR non-eligible) at study day 1 (population at risk = 0)

Show code

Show code

**Table A1:**
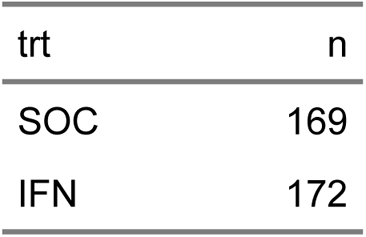
Starting number of households per treatment arm.

| trt | n |
| --- | --- |
| SOC | 169 |
| IFN | 172 |

Show code

**Table A2:** Breakdown of corresponding participants.

| trt | IC | CEP | CE | NE |
| --- | --- | --- | --- | --- |
| SOC | 169 | 81 | 165 | 150 |
| IFN | 172 | 80 | 177 | 178 |

Show code

**Table A3:** Number of households with a PCR positive index case.

| trt | n |
| --- | --- |
| SOC | 149 |
| IFN | 159 |

Show code

**Table A4:** Breakdown of corresponding participants.

| trt | IC | CEP | CE | NE |
| --- | --- | --- | --- | --- |
| SOC | 149 | 77 | 142 | 134 |
| IFN | 159 | 76 | 164 | 164 |

Show code

**Table A5:** Households with a PCR positive index case and population at risk > 0 at start of period 1.

| trt | n |
| --- | --- |
| SOC | 123 |
| IFN | 136 |

**Analysis is carried out using this latter number of households.**

Show code

### Analysis

Show code

**Table A6:**
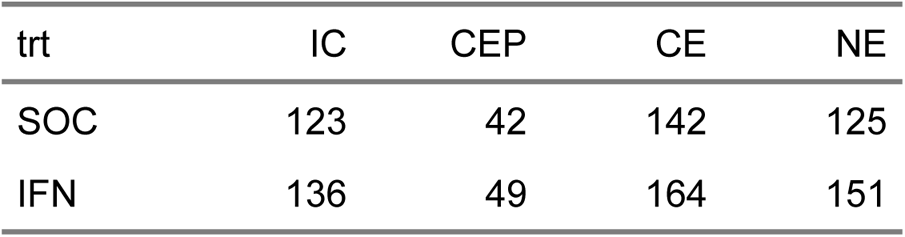
Breakdown of corresponding participants.

While **eligible** contacts (CE) that were PCR(+) at the start of the study are tabulated separately above (as CEP), **ineligible** contacts (NE) that were PCR(+) at the start of the study were not explicitly identified in the same manner. Our analysis considers only the at risk population (at the start of each period), so we further exclude these participants below.

Show code

**Table A7:** Analysis population: Household contact population excluding those that were PCR(+) on study day 1.

| trt | CE | NE |
| --- | --- | --- |
| SOC | 142 | 104 |
| IFN | 164 | 129 |

### Incidence of infection

**Finally**, in order to provide some context, we report the incidence of infection in each study arm during the first period (on treatment; D1-11 inclusive), for eligible and non-eligible household contact populations.

Show code

**Table A8:**
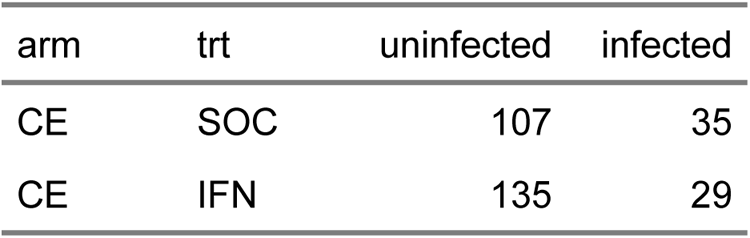
Incidence of infection in the eligible household contact population during treatment.

| arm | trt | uninfected | infected |
| --- | --- | --- | --- |
| CE | SOC | 107 | 35 |
| CE | IFN | 135 | 29 |

Show code

**Table A9:** Incidence of infection in the non-eligible household contact population during treatment.

| arm | trt | uninfected | infected |
| --- | --- | --- | --- |
| NE | SOC | 79 | 25 |
| NE | IFN | 103 | 26 |

### Model 2 vs. Model 1

This comparison answers the following question:

Is treatment with IFN associated with a reduction in the number of incident positive saliva PCR in household contacts, adjusting for individual risk factors (age, sex, and treatment eligibility) and household risk factors (num. individuals in household, num. vaccinated individuals, num. infected at start of period, viral load of index case), as well as treatment period (D1-11 vs. D11-29)?

Show code

| term | npar | AIC | BIC | logLik | deviance | statistic | df | p.value |
| --- | --- | --- | --- | --- | --- | --- | --- | --- |
| mod0 | 12.000 | 622.676 | 681.117 | -299.338 | 598.676 |  |  |  |
| mod1 | 13.000 | 620.136 | 683.446 | -297.068 | 594.136 | 4.541 | 1.000 | 0.033 |

Show code

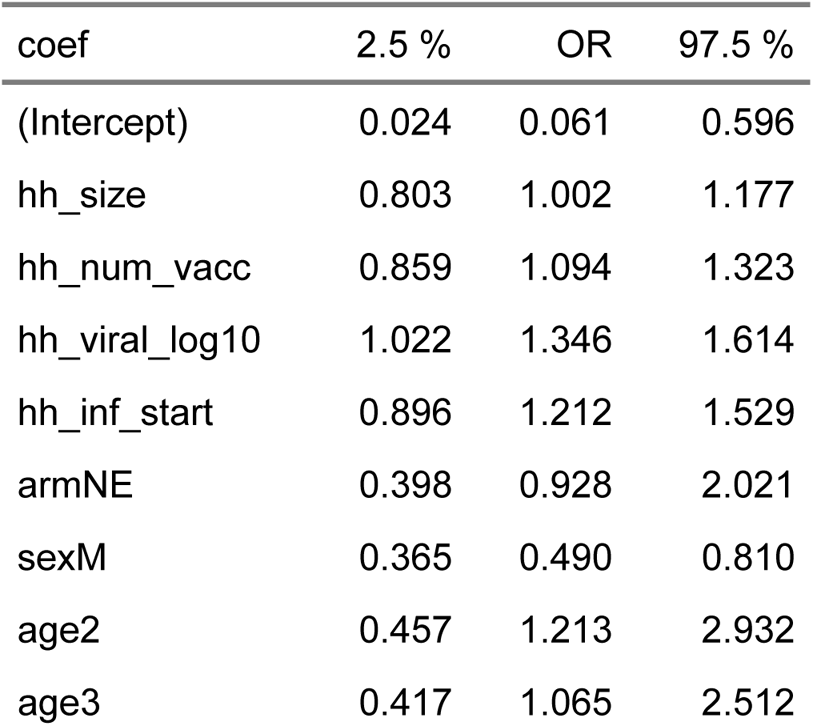
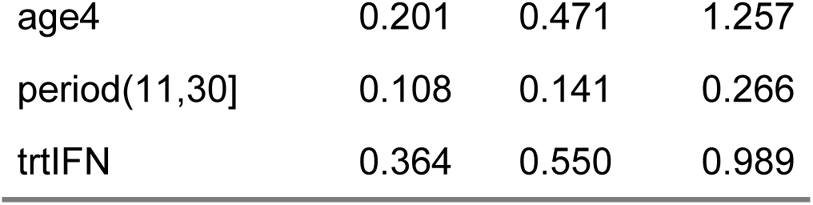

### Conclusion

**Treatment with IFN is associated with a significant reduction in the odds of a positive saliva PCR for all household contacts compared to standard of care (p = 0.033; OR = 0.550, 95% CI = 0.364 to 0.989).**

### Model 3 vs. Model 2

This comparison answers the following question:

Does the effect of treatment on the number of incident positive saliva PCR in household contacts change between study periods?

Show code

| term | npar | AIC | BIC | logLik | deviance | statistic | df | p.value |
| --- | --- | --- | --- | --- | --- | --- | --- | --- |
| mod0 | 12.000 | 618.151 | 676.592 | -297.076 | 594.151 |  |  |  |
| mod1 | 13.000 | 619.077 | 682.387 | -296.538 | 593.077 | 1.075 | 1.000 | 0.300 |

Show code

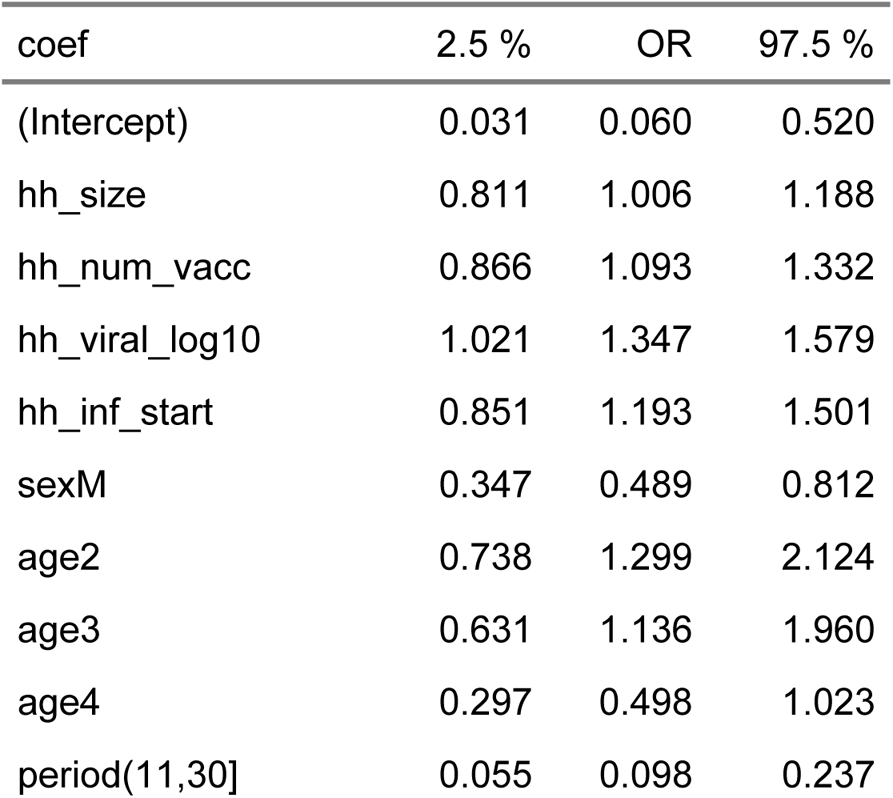
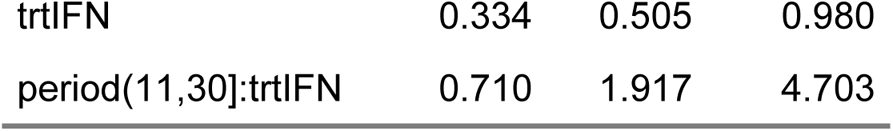

### Conclusion

**Inconclusive: the effect of treatment does not appear to change between period 1 and period 2 (p = 0.300).**

Show code

**The point estimate for the treatment effect in the second period is OR = 0.968 (95% CI = 0.405 to 2.840) compared to OR = 0.550 (95% CI = 0.364 to 0.989) in the first period.**

### Summary Statement

In this ***exploratory*** analysis of the PCR results during periods 1 and 2 using the larger HC study population (eligible plus ineligible household contacts), and additionally considering treatment periods (during treatment, D1-D11; after treatment, D12-D29), IFN given to treatment eligible household contacts (and the COVID(+) index case) significantly reduced the odds of positive PCR results for all (non-positive at study day 1) household contacts (IFN likelihood ratio test p = 0.033; OR = 0.550, 95% CI = 0.364 to 0.989). Positive PCR tests are significantly less common in the second period (OR = 0.141, 95% CI = 0.108 to 0.266). The results for treatment and period summarized above were taken from the second model which excluded the treatment by period interaction as the interaction term was not significant (interaction LRT p = 0.300).

### Figures

**Figure A1:**
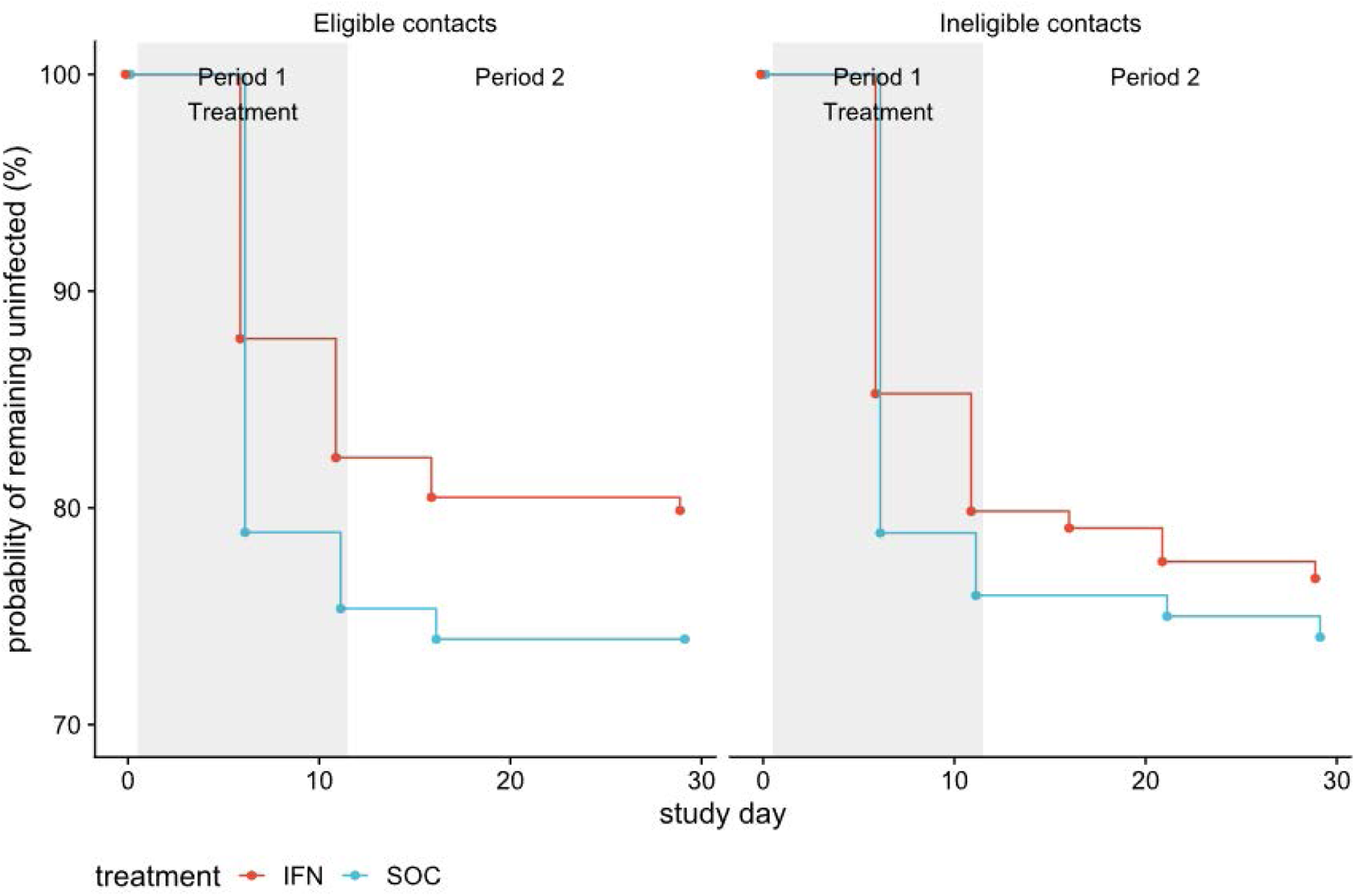
Treatment eligible (left) or ineligible (right) household contact population at risk over time stratified by randomization (red, treatment [IFN]; blue, standard of care).

## References

1. Alizon S, Haim-Boukobza S, Foulongne V, et al. Rapid spread of the SARS-CoV-2 Delta variant in some French regions, June 2021. Euro Surveill 2021;26(28). DOI: 10.2807/1560-7917.ES.2021.26.28.2100573.

2. Mishra S, Mindermann S, Sharma M, et al. Changing composition of SARS-CoV-2 lineages and rise of Delta variant in England. EClinicalMedicine 2021;39:101064. DOI: 10.1016/j.eclinm.2021.101064.

3. Twohig K, Nyberg T, Zaidi A, et al. Hospital admission and emergency care attendance risk for SARS-CoV-2 delta (B.1.617.2) compared with alpha (B.1.1.7) variants of concern: a cohort study. Lancet Infectious Diseases 2021. DOI: 10.1016/S1473-3099(21)00475-8.

4. Winger A, Caspari T. The Spike of Concern-The Novel Variants of SARS-CoV-2. Viruses 2021;13(6). DOI: 10.3390/v13061002.

5. Lyngse FP, Mortensen LH, Denwood MJ, et al. SARS-CoV-2 Omicron VOC Transmission in Danish Households. medRxiv 2021.

6. Nyberg T, Ferguson NM, Nash SG, et al. Comparative analysis of the risks of hospitalisation and death associated with SARS-CoV-2 omicron (B.1.1.529) and delta (B.1.617.2) variants in England: a cohort study. Lancet 2022;399(10332):1303–1312. DOI: 10.1016/S0140-6736(22)00462-7.

7. Le Rutte E, Shattock A, Chitnis N, Kelly SL, Penny M. Assessing impact of Omicron on SARS-CoV-2 dynamics and public health burden. medRXiv. Online server: Cold Spring Harbor Laboratory; 2021.

8. Centers for Disease Control and Prevention. Implementation of Mitigation Strategies for Communities with Local COVID-19 Transmission. CDC. May 23 2021.

9. Milne GJ, Xie S, Poklepovich D, O’Halloran D, Yap M, Whyatt D. A modelling analysis of the effectiveness of second wave COVID-19 response strategies in Australia. Sci Rep 2021;11(1):11958. DOI: 10.1038/s41598-021-91418-6.

10. National Incident Room Surveillance Team. COVID-19 Australia: Epidemiology Report 38 Reporting period ending 28 March 2021. Commun Dis Intell (2018) 2021;45. DOI: 10.33321/cdi.2021.45.19.

11. Houvessou GM, Souza TP, Silveira MFD. Lockdown-type containment measures for COVID-19 prevention and control: a descriptive ecological study with data from South Africa, Germany, Brazil, Spain, United States, Italy and New Zealand, February - August 2020. Epidemiol Serv Saude 2021;30(1):e2020513. DOI: 10.1590/S1679-49742021000100025.

12. Conway SR, Lazarski CA, Field NE, et al. SARS-CoV-2-Specific T Cell Responses Are Stronger in Children With Multisystem Inflammatory Syndrome Compared to Children With Uncomplicated SARS-CoV-2 Infection. Front Immunol 2021;12:793197. DOI: 10.3389/fimmu.2021.793197.

13. Cutler DM, Summers LH. The COVID-19 Pandemic and the $16 Trillion Virus. JAMA 2020;324(15):1495–1496. DOI: 10.1001/jama.2020.19759.

14. Verschuur J, Koks EE, Hall JW. Global economic impacts of COVID-19 lockdown measures stand out in high-frequency shipping data. PLoS One 2021;16(4):e0248818. DOI: 10.1371/journal.pone.0248818.

15. Chemen S, Gopalla YN. Lived experiences of older adults living in the community during the COVID-19 lockdown - The case of mauritius. J Aging Stud 2021;57:100932. DOI: 10.1016/j.jaging.2021.100932.

16. Mamun MA. Suicide and Suicidal Behaviors in the Context of COVID-19 Pandemic in Bangladesh: A Systematic Review. Psychol Res Behav Manag 2021;14:695–704. DOI: 10.2147/PRBM.S315760.

17. Callaway E. Beyond Omicron: what’s next for COVID’s viral evolution. Nature 2021;600(7888):204–207. DOI: 10.1038/d41586-021-03619-8.

18. Rahmani H, Davoudi-Monfared E, Nourian A, et al. Interferon β-1b in treatment of severe COVID-19: A randomized clinical trial. Int Immunopharmacol 2020;88:106903. (In eng). DOI: 10.1016/j.intimp.2020.106903.

19. Alavi Darazam I, Shokouhi S, Pourhoseingholi MA, et al. Role of interferon therapy in severe COVID-19: the COVIFERON randomized controlled trial. Sci Rep 2021;11(1):8059. (In eng). DOI: 10.1038/s41598-021-86859-y.

20. Davoudi-Monfared E, Rahmani H, Khalili H, et al. A Randomized Clinical Trial of the Efficacy and Safety of Interferon β-1a in Treatment of Severe COVID-19. Antimicrob Agents Chemother 2020;64(9) (In eng). DOI: 10.1128/aac.01061-20.

21. Feld JJ, Kandel C, Biondi MJ, et al. Peginterferon lambda for the treatment of outpatients with COVID-19: a phase 2, placebo-controlled randomised trial. Lancet Respir Med 2021;9(5):498–510. (In eng). DOI: 10.1016/s2213-2600(20)30566-x.

22. Meng Z, Wang T, Chen L, et al. The Effect of Recombinant Human Interferon Alpha Nasal Drops to Prevent COVID-19 Pneumonia for Medical Staff in an Epidemic Area. Curr Top Med Chem 2021;21(10):920–927. (In eng). DOI: 10.2174/1568026621666210429083050.

23. National Institutes of Health. Covid-19 Treatment Guidelines. In: Health NIH, ed.: United States Goverment; 2021.

24. Lee AJ, Ashkar AA. The Dual Nature of Type I and Type II Interferons. Front Immunol 2018;9:2061. DOI: 10.3389/fimmu.2018.02061.

25. Mazewski C, Perez RE, Fish EN, Platanias LC. Type I Interferon (IFN)-Regulated Activation of Canonical and Non-Canonical Signaling Pathways. Front Immunol 2020;11:606456. DOI: 10.3389/fimmu.2020.606456.

26. Wang BX, Fish EN. Global virus outbreaks: Interferons as 1st responders. Semin Immunol 2019;43:101300. DOI: 10.1016/j.smim.2019.101300.

27. World Health Organization. Informal consultation on the role of therapeutics in COVID-19 prophylaxis and post-exposure prophylaxis. . Worl Health Organization.

28. Gentile I, Maraolo AE, Piscitelli P, Colao A. COVID-19: Time for Post-Exposure Prophylaxis? Int J Environ Res Public Health 2020;17(11):3997. (In eng). DOI: 10.3390/ijerph17113997.

29. Iturriaga C, Eiffler N, Aniba R, et al. A cluster randomized trial of interferon ss-1a for the reduction of transmission of SARS-Cov-2: protocol for the Containing Coronavirus Disease 19 trial (ConCorD-19). BMC Infect Dis 2021;21(1):814. DOI: 10.1186/s12879-021-06519-4.

30. World Health Organization. Household transmission investigation protocol for 2019-novel coronavirus (COVID-19) infection. Epidemiological protocol. Geneva: World Health Organization, February 28th 2020 2020. (WHO/2019-nCoV/HHtransmission/2020.4) (https://www.who.int/publications/i/item/household-transmission-investigation-protocol-for-2019-novel-coronavirus-(2019-ncov)-infection).

31. Reess J, Haas J, Gabriel K, Fuhlrott A, Fiola M. Both paracetamol and ibuprofen are equally effective in managing flu-like symptoms in relapsing-remitting multiple sclerosis patients during interferon beta-1a (AVONEX) therapy. Mult Scler 2002;8(1):15–8. DOI: 10.1191/1352458502ms771sr.

32. National Health and Medical Research Council. Guidance: Safety monitoring and reporting in clinical trials involving therapeutic goods. Canberra: National Health and Medical Research Council; 2016.

33. Hofmeijer J, Anema PC, van der Tweel I. New algorithm for treatment allocation reduced selection bias and loss of power in small trials. J Clin Epidemiol 2008;61(2):119–24. DOI: 10.1016/j.jclinepi.2007.04.002.

34. National Institute of Statistics. Chile Population and Housing Census 2017. Santiago, Chile: National Institute of Statistics (Chile), 2021. (http://ghdx.healthdata.org/record/chile-population-and-housing-census-2017).

35. World Health Organisation. Situation report - 50. Coronavirus disease 2019 (COVID-19) 50. World Health Organisation, 2020. (https://www.who.int/emergencies/diseases/novel-coronavirus-2019/situation-reports).

36. Yi B, Fen G, Cao D, et al. Epidemiological and clinical characteristics of 214 families with COVID-19 in Wuhan, China. Int J Infect Dis 2021. DOI: 10.1016/j.ijid.2021.02.021.

37. Curmei M, Ilyas A, Evans O, Steinhardt J. Constructing and adjusting estimates for household transmission of SARS-CoV-2 from prior studies, widespread-testing and contact-tracing data. Int J Epidemiol 2021;50(5):1444–1457. DOI: 10.1093/ije/dyab108.

38. Bates DM, M.; Bolker, B.; Walker, S. Fitting linear mixed-effects models using lme4. Journal of Statistical Software 2015;67(1):1–48. DOI: doi:10.18637/jss.v067.i01.

39. Goodrich B, Gabry J, Ali I, S. B. Bayesian applied regression modeling via Stan. R package version 2.21.1. rstanarm 2020 (https://mc-stan.org/rstanarm.).

40. Brilleman SL, Crowther MJ, Moreno-Betancur M, Buros Novik J, Wolfe R. Joint longitudinal and time-to-event models via Stan. StanCon 2018. 10-12 Jan 2018. Pacific Grove C, USA. https://github.com/stan-dev/stancon_talks/. Joint longitudinal and time-to-event models via Stan. StanCon 2018. Pacific Grove, CA, USA2018.

41. Hu X, Seddighzadeh A, Stecher S, et al. Pharmacokinetics, pharmacodynamics, and safety of peginterferon beta-1a in subjects with normal or impaired renal function. J Clin Pharmacol 2015;55(2):179–88. DOI: 10.1002/jcph.390.

42. Labhardt ND, Smit M, Petignat I, et al. Post-exposure Lopinavir-Ritonavir Prophylaxis versus Surveillance for Individuals Exposed to SARS-CoV-2: The COPEP Pragmatic Open-Label, Cluster Randomized Trial. EClinicalMedicine 2021;42:101188. DOI: 10.1016/j.eclinm.2021.101188.

43. Ben K, Alex M, Mariya K, et al. Nature Portfolio 2022. DOI: 10.21203/rs.3.rs-1121993/v1.

## Supplementary References

1. Goodrich B GJ, Ali I, Brilleman S. rstanarm: Bayesian applied regression modeling via Stan. R package version 2.21.3 ed2022.

